# The benefits and harms of cancer screening programmes for adults with intellectual disabilities: A systematic review

**DOI:** 10.1101/2025.08.01.25332659

**Authors:** Martin McMahon, Samantha Flynn, Samantha A Johnson, Chris Stinton

## Abstract

**Introduction:** Cancer screening programmes are an important public health initiative aimed at reducing morbidity and mortality through the early detection of breast, cervical, and colorectal cancers. The available evidence points towards lower screening participation in cancer screening programmes among people with intellectual disabilities, but the balance of benefits and harms of screening is unknown. The aim of this systematic review is to examine the health outcomes (mortality, morbidity) and harms of cancer screening programmes for adults with intellectual disabilities.

**Materials and methods:** The review is registered on the Open Science Framework Registries: https://osf.io/8vmkb. A systematic search of published peer-reviewed and grey literature was conducted from inception up to 28^th^ February 2025. MEDLINE, EMBASE, Web of Science, PsycINFO, and relevant organisational websites were searched. Additionally, experts in the field were consulted about evidence on the health outcomes and harms of national cancer screening programmes for adults with intellectual disabilities. Two reviewers independently screened titles and abstracts and assessed full texts against the eligibility criteria.

**Results:** A total of 3,104 records were identified, and 232 full-text articles were assessed for eligibility. No study met the inclusion criteria.

**Conclusions:** There is a critical lack of evidence regarding the benefits and harms of cancer screening programmes for people with intellectual disabilities. There are numerous studies relating to coverage, uptake, and factors influencing participation in cancer screening programmes. Participation should not be assumed to equate to better outcomes, and there is a real risk of ‘equity-washing’ if this is used as a basis for practice. There is, therefore, an urgent need to examine the full screening pathway continuum for people with intellectual disabilities, along with the associated outcomes, benefits, and harms.

## Introduction

Cancer is a major societal, public health, and economic problem, ranking as a leading cause of death worldwide (1). Cancer screening programmes, which detect pre-cancerous or early-stage cancer in asymptomatic individuals are an important public health initiative that increases the chance of survival (2). However, screening is also associated with harms (3) such as discomfort and future health risks (e.g. radiation exposure)(4) from tests, inaccurate results that cause false reassurance (false negative test results)(5) or anxiety(6, 7) (false positive test results)(8), and overdiagnosis(9–11) (the detection of disease that would not have caused symptoms during a person’s lifetime) which leads to unnecessary overtreatment. While these are not always avoidable, in an effective screening programme the benefits will outweigh the harms.

Most cancer screening programmes focus on detecting cancers of the bowel, female breast, and cervix, and these cancers contribute substantially to the overall cancer burden (12). Internationally, there is significant variability in how screening programmes are delivered, with non-uniformed approaches likely to mask differences, making like-for-like comparisons difficult. Western European countries typically experience the highest rates of uptake and coverage of cancer screening programmes,(13) with socioeconomic status and social vulnerability factors (14) being known to negatively influence uptake and outcomes (15).

Cancer screening uptake and coverage in people with intellectual disabilities have been reported as being lower than that of the general population (16–19). Increased social vulnerability factors, health inequalities, and inequities regarding access and accessibility are frequently cited as issues that impact healthcare utilisation (20, 21). However, it is not fully clear how these factors impact the uptake of national cancer screening programmes in their entirety, though the mechanisms are likely similar (22). Despite dying younger, mortality ages for people with intellectual disabilities are increasing, meaning they are more likely to be exposed to the largest risk factor for developing cancers—aging (23, 24).Emerging evidence from Canada (25, 26), Scotland (27), and England (28) highlights that some cancers for which screening programmes exist appear to be occurring at a younger age in people with intellectual disabilities population. Moreover, when these cancers are detected they are often identified at later stages than the general population (26). This limits the chance of having curative treatment. That such inequities exist provides a concerning picture regarding the detection of cancers where population-level interventions exist but do not meet the needs of people with intellectual disabilities.

Screening programmes recognise the need for a more inclusive approach towards screening, with the Irish national screening service developing an equity framework in 2023 (29) and Public Health England developing their screening inequalities strategy in 2020 (30) to better understand and improve equity in cancer screening. Nevertheless, barriers to screening continue to exist for people with intellectual disabilities (20). In parallel with the general literature, it is likely that these barriers follow a health equity gradient influenced by broader general socioeconomic and environmental conditions where people from more marginalised and hard-to-reach groups are more likely to be influenced by the negative factors that also influence the determinants of health and well-being (31–33).

Due to the existing challenges and complex health needs of people with intellectual disabilities, it is important to understand participation in national cancer screening programmes. No attempt has yet been made to synthesise the existing published evidence exploring the relative harms and benefits of cancer screening for people with intellectual disabilities. Consequently, this review aims to evaluate the health outcomes (e.g. mortality, morbidity) of participation in national cancer screening programmes for people with intellectual disabilities. In doing so, this systematic review aims to provide evidence to help understand the benefits and limitations for this population by identifying gaps and areas for improvement and future practice.

### Research questions

1. What are the health outcomes (e.g. mortality, morbidity) of cancer screening programmes for adults who have intellectual disabilities?
2. What are the harms of cancer screening programmes for adults who have intellectual disabilities?

## Materials and methods

The review is registered on the Open Science Framework Registries: https://osf.io/8vmkb

### Identification and selection of studies

#### Search strategy

We undertook a comprehensive search strategy comprising: (1) searches of electronic bibliographic databases; (2) searches of websites of intellectual disabilities charities and cancer charities; (3) contacting known experts in the field and through discussions during an international workshop in May 2025; and (4) scrutiny of references of included studies and relevant systematic reviews.

The electronic bibliographic databases search strategy was developed in MEDLINE (Ovid) using terms relating to (1) intellectual disability, (2) cancer, and (3) screening, and was adapted for EMBASE (Ovid), Web of Science (Clarivate), and PsycInfo (Ovid). Copies of the full search strategies are provided in Supplementary file 1. Searches were conducted on the 28^th^ February 2025.

To identify additional/unpublished research, we searched the websites of a sample of charitable organisations that support either (1) people who have intellectual disabilities, or (2) people who have bowel, breast, or cervical cancer and provide research grants. The organisations were either known to the review team or identified through a Google search (“national [breast/bowel/cervical] cancer charities grant funding research”), with relevant returns from the first two search results pages being searched. A list of the organisations searched is provided in Supplementary file 2. Each organisation’s website was searched by entering the term “screening” (intellectual disability charities) or “disability” (cancer charities) into the website’s search box. Searches were conducted from the 7^th^ to the 13^th^ March 2025.

### Study eligibility criteria

Studies reporting quantitative data, that were published in the English language, and investigated the health benefits (e.g., mortality, morbidity) and harms of population or targeted bowel, breast, and cervical cancer screening programmes for adults who have intellectual disabilities were eligible for inclusion in this review. We excluded studies of children and young people (aged 17 and under), people who did not have an intellectual disability, and where more than 10% of the study sample did not meet our inclusion criteria. Studies where the method of cancer detection was opportunistic screening, case-finding, one-off testing outside of an organised screening programme or routine examinations were also excluded, as were studies that provided insufficient information for assessment of methodological quality/risk of bias to be conducted, or no numerical outcome data.

Documents not available in the English language, studies where it could not be determined if the inclusion criteria were met, and literature reviews were also not eligible for inclusion in this review.

### Review strategy

Study selection was conducted through the Covidence platform (34) for the electronic databases, and directly through the websites of cancer and intellectual disabilities organisations. For both electronic database and website searches, two reviewers independently assessed titles and abstracts. Any paper considered potentially relevant by either reviewer was retained for full text assessment. Full text assessment was conducted independently by two reviewers against the review eligibility criteria, with any disagreements resolved by consensus or through discussion with a third reviewer.

## Results

### Searching, sifting, and sorting

Full details of the flow of studies through the review are reported in Figure 1. We identified 2,262 unique records through database searches and 842 documents through the websites of cancer and intellectual disabilities organisations. After examination of titles and abstracts, 232 documents were retained for full text assessment (electronic databases: n = 231; websites: n = 1). None met the inclusion criteria for this review. Appendix 3 lists the studies that were excluded at the full text assessment stage of this review.

**Figure 1.**
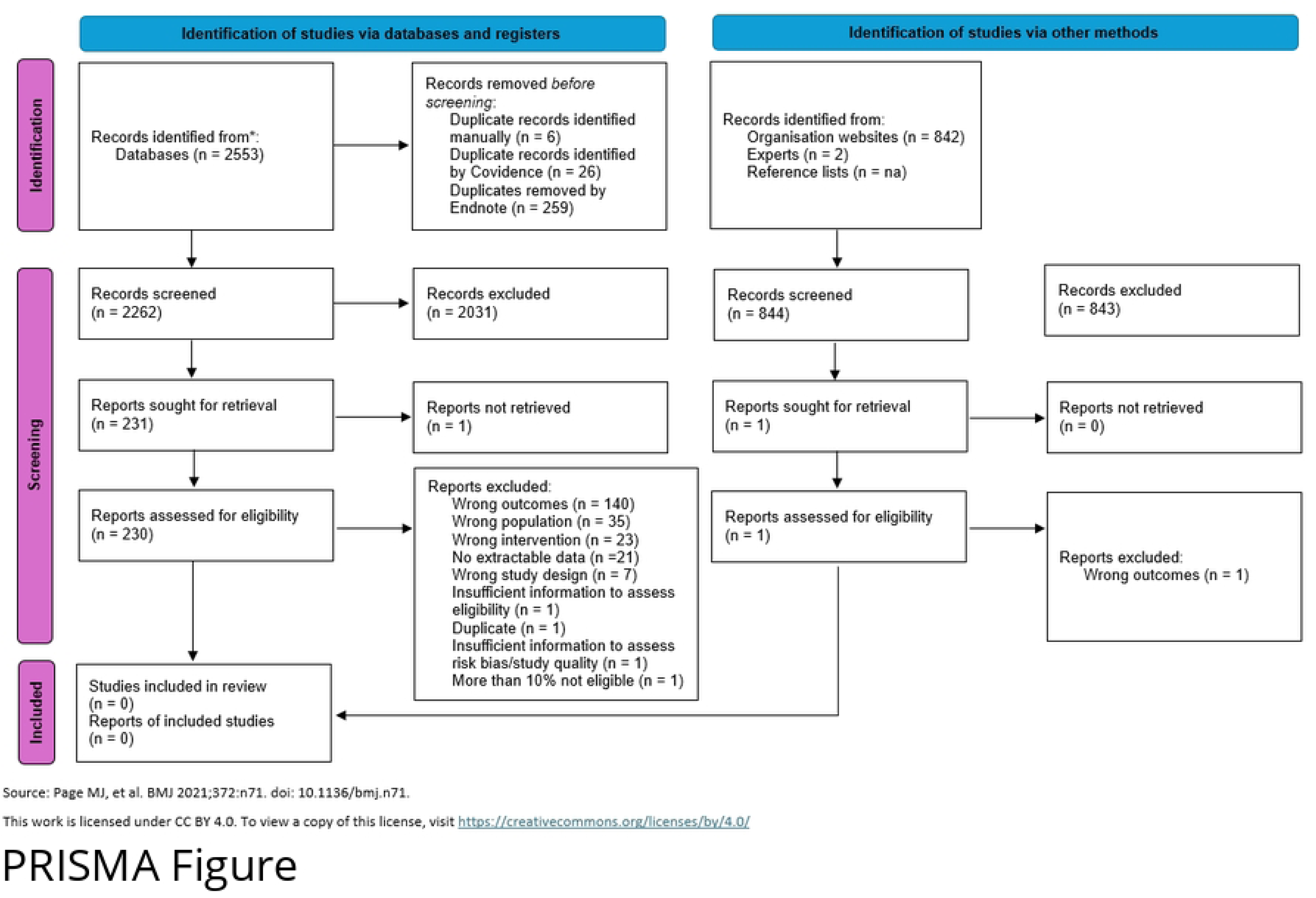
PRISMA flow diagram

## Discussion

In this systematic review, we intended to examine the health outcomes (e.g. mortality, morbidity) and harms of screening for breast, bowel, and cervical cancers for people with intellectual disabilities. Despite extensive searches of peer-reviewed and grey literature, we did not identify any studies that met the review eligibility criteria. The published evidence in this field broadly relates to the uptake and determinants of cancer screening programmes (16–20). While this is an important consideration, it does not address the overarching aims and objectives of screening, which are to improve morbidity and mortality through the early detection of disease (2). A significant evidence gap exists regarding health outcomes from cancer screening programmes for people with intellectual disabilities, including its relative benefits and harms. Therefore, recommendations from existing research that advocate for increased uptake and coverage need to be fully considered in terms of their overall health benefits for this population.

Cancer screening programmes offer substantial benefits, particularly with respect to mortality reduction from cervical(35) and colorectal screening (36), which may provide a greater net benefit than breast screening (37). For people with intellectual disabilities, the screening process needs to be understood as a continuum, not merely as a single event focused on coverage and uptake. While there is growing evidence in this population around invitation and attendance, greater emphasis must also be placed on exploring health outcomes, test performance, diagnostic follow-up, treatment initiation, positive outcomes, and harms, which are the fundamental to screening (2). Solely focusing on coverage and uptake risks ‘equity-washing’, whereby numerical participation is improved without knowing whether the programmes actually enhance health outcomes for people with intellectual disabilities. This lack of evidence is particularly concerning given the atypical health presentations in this population, including different baselines for life expectancy and increased multimorbidity (24, 38–40).

People with intellectual disabilities face unique risks related to the capacity to consent, physiological differences, and the invasiveness and validity of some tests (41, 42). For example, in the Netherlands, Banda and colleagues(19) reported higher rates of invalid and inconclusive test results in the breast cancer, cervical cancer, and colon cancer screening programmes among people with intellectual disabilities compared with the general population. Although not reported in the study, these findings may point towards potential harms of screening, particularly increased psychological and physical burden, but further research is needed. Additionally, the experience of screening and its psychological impacts should not be overlooked for this population, who often experience difficulties with self-reporting and health literacy (43, 44). A recent systematic review of general asymptomatic populations reported that while most people view screening positively, those who receive false-positive results often experience increased levels of anxiety (7). A separate study on mammography found that women were less likely to return for future screenings following false-positive results, particularly when short-interval follow-up or biopsy was recommended, raising concerns about ongoing engagement in routine screening (45).

Colorectal screening may offer clear benefits for people with intellectual disabilities, given the high prevalence of these cancers in this population (27, 46). However, when positive results from screening tests (e.g., Faecal Immunochemical Test) or presenting symptoms necessitate further investigation, colonoscopy is typically required and often involves heavy sedation and/or general anaesthesia. Such procedures introduces increased risks for people with intellectual disabilities and require appropriate adjustments (47). These harm–benefit trade-offs must be appropriately assessed and considered but, as the lack of results in our systematic review demonstrate, there is no current evidence to support people with intellectual disabilities to make decisions about whether to participate in cancer screening programmes.

An recent example of the importance of understanding harm–benefit trade-offs come from a modelling study simulating the benefits and harms of annual, biennial, triennial, and one-time digital mammography screening in women with Down syndrome(48). In this study, screening was assessed to be less favourable for women with Down syndrome compared to average-risk women without Down syndrome, due to their lower baseline risk of breast cancer and reduced life expectancy. While the study was based on a hypothetical cohort, it does highlight that standard screening guidelines developed for average-risk populations may be deficient, or even inappropriate, for people with Down syndrome.

In this context, similar patterns of inequity and have been observed in other underserved groups highlighting the need to understand health outcomes and understanding the benefits and harm of cancer screening programmes. Cancer screening participation rates are reported to be significantly lower among African American, Asian American, Latinx American, and American Indian/Alaska Native populations (49) as well as among transgender and gender-nonconforming adults (50), LGBTQ+ populations (51), and individuals with severe mental illness. (52) While much of the evidence also focuses on coverage and uptake, there is growing documentation of downstream harms, such as poorer cancer outcomes among African Americans (53), and people with mental illness (54). Similar inequalities and inequities in cancer risk and outcomes are now being observed in the intellectual disability population (28). For example, Heslop et al. (2022) found that 43% of adults diagnosed with colorectal cancer in England were aged 18–59 years, suggesting that the age threshold for colorectal screening (over 60 years) in people with intellectual disabilities may be inequitable.

An example of screening policy being altered to meet the needs of population subgroups comes from the UK cervical screening programme where self-sampling was recently recommended as an alternative to clinician-sampling for under-screened women (55). This example illustrates how recommendations evolve to reflect population-specific risk profiles.

As a priority, we argue for an evaluation of the entire screening pathway for people with intellectual disabilities to determine where processes succeed and where they fall short for this population. This should include assessments of health outcomes, economic efficiency, and benefit–harm balance. Without this evidence, there is a risk that efforts to improve uptake may be interpreted as improvements in outcomes, when they may, in fact, lead to unintended harm. We therefore assert that, collectively, we must explore and understand what happens after people with intellectual disabilities participate in screening, particularly in terms of the balance of benefits and harms. Only then can we consider reasonable adjustments to enhance health outcomes for people with intellectual disabilities when accessing cancer screening.

### Strengths and limitations

To our knowledge, this is the first systematic review to examine the benefits and harms of cancer screening programmes for people who have intellectual disabilities. We followed standard methodology for conducting systematic reviews, and the protocol is publicly available on the Open Science Framework (https://osf.io/8vmkb). We searched a broad range of sources for evidence (electronic databases, grey literature in the form of relevant organisation websites, contact with experts), without imposing limits on study design beyond a necessity for studies to present some relevant quantitative data.

Our review has several limitations. First, we did not involve people with intellectual disabilities in this review. This has implications for the applicability of the review as while mortality, morbidity, and harms are essential outcomes of screening programmes they are not necessarily the only outcomes of interest for people with intellectual disabilities themselves. Second, we took a pragmatic approach to determining which websites to search for relevant research. While this is likely to capture the websites of the largest organisations relating to cancer and intellectual disabilities, the results returned are determined by the algorithm used for that search engine; different organisation websites might have been identified by other search engines. There are also numerous other sources of grey literature that we did not explore (e.g., doctoral theses). Finally, we did not search for studies in languages other than English. It is possible that relevant studies in languages other than English could have been missed.

### Future Research

This review highlights a significant gap in the evidence relating to health outcomes from cancer screening programmes for people with intellectual disabilities. Existing research has largely focused on coverage and uptake, but not on the benefits and harms associated with cancer screening for this population. From this perspective, research should view screening programmes as a continuum and, using longitudinal data, map the full trajectory from invitation to participation, diagnosis, and outcomes for people with intellectual disabilities. This would provide a greater understanding of the health outcomes of screening for this population.

Additionally, it be important to estimate the benefit–harm ratio and cost-effectiveness of screening strategies for this population, accounting for different scenarios in terms of this population’s life expectancy and associated comorbidities. It is also important to consider the lived experiences of people with intellectual disabilities, as well as their families and supporters, in relation to participating in cancer screening programmes. This is particularly important to help understand the and psychological impacts of cancer screening programmes.

Overall, it is critically important that future research addresses the potential for ‘equity-washing’, where increases in coverage and uptake are misrepresented as improved outcomes, without consideration of the actual benefit–harm balance.

## Conclusion

In summary, this systematic review highlights a substantial evidence gap concerning the benefits and harms of cancer screening programmes for adults with intellectual disabilities. We found no studies that had evaluated whether cancer screening improves outcomes or causes harm in people with intellectual disabilities. Without this evidence and knowledge of downstream effects, the balance of benefits and harms of cancer screening in people with intellectual disabilities is unknown. ancer screening must be assessed as a continuum of care, not solely through measures of uptake and coverage.

## Data Availability

All relevant data are within the manuscript and its Supporting Information files.

## Acknowledgement

The views expressed in this publication are those of the authors and do not necessarily reflect those of our host institutions or any funders.

## Supporting information

## Author contributions

### CRediT authorship contribution statement

**Martin McMahon:** Conceptualisation, methodology, validation, investigation, writing - original draft, writing - review and editing, project administration

**Samantha Flynn:** Conceptualisation, methodology, validation, investigation, writing - review and editing

**Samantha A Johnson:** Methodology, investigation, writing - review and editing,

**Chris Stinton:** Conceptualisation, methodology, validation, investigation, writing - original draft, writing - review and editing, supervision

## Appendix 1. Database searches

| Medline ALL (OVID) 1946- |  |  |
| --- | --- | --- |
| # | Search terms | Results |
| 1. | ((intellectual or learning or neurodevelop* or development* or mental) adj4 (disabilit* or impair* or handicap or retard* or deficient*)).mp | 175422 |
| 2. | exp Intellectual Disability/ | 109278 |
| 3. | exp Learning Disabilities/ | 24548 |
| 4. | exp developmental disabilities/ | 23316 |
| 5. | Neurodevelopmental Disorders/ | 5771 |
| 6. | 1 or 2 or 3 or 4 or 5 | 227553 |
| 7. | (cancer* or neoplasm* or carcinoma* or tumor* or tumour* or oncolog*).mp | 4514628 |
| 8. | (colorectal or bowel or breast or mammar* or cervix or cervic*).mp | 1248077 |
| 9. | 7 and 8 | 816495 |
| 10. | exp Colorectal Neoplasms/ | 253340 |
| 11. | exp Breast Neoplasms/ | 364569 |
| 12. | exp Uterine Cervical Neoplasms/ | 89875 |
| 13. | 9 or 10 or 11 or 12 | 910347 |
| 14. | ((((fecal or faecal) and test) or occult blood test or ((fecal or faecal) and immunochemical test*) or (colonoscop* or endoscop* or sigmoid*) or mammogra* or ((human and (papillomavirus or papilloma virus)) or HPV))).mp | 443116 |
| 15. | exp Occult Blood/ | 6652 |
| 16. | exp Colonoscopy/ | 36825 |
| 17. | exp Endoscopy/ | 427442 |
| 18. | exp Sigmoidoscopy/ | 4928 |
| 19. | exp Mammography/ | 34820 |
| 20. | (screen* or early detect*).mp | 1063010 |
| 21. | exp Mass Screening/ | 149709 |
| 22. | exp "Early Detection of Cancer"/ | 43128 |
| 23. | 14 or 15 or 16 or 17 or 18 or 19 or 20 or 21 or 22 | 1680360 |
| 24. | 6 and 13 and 23 | 212 |
| 25. | limit 24 to English language | 206 |

| Embase Classic + Embase (OVID), 1947- |  |  |
| --- | --- | --- |
| # | Search terms | Results |
| 1. | ((intellectual or learning or neurodevelop* or development* or mental)<br>adj4 (disabilit* or impair* or handicap or retard* or deficient*).mp | 222662 |
| 2. | intellectual disabilit*.mp. or exp intellectual impairment/ | 681513 |
| 3. | learning disabilit*.mp. or exp learning disorder/ | 46894 |
| 4. | developmental disabilit*.mp. or exp developmental disorder/ | 72178 |
| 5. | (neurodevelopmental disabilit* or neurodevelopmental disorder*).mp | 29092 |
| 6. | 1 or 2 or 3 or 4 or 5 | 855557 |
| 7. | (cancer* or neoplasm* or carcinoma* or tumor* or tumour* or oncolog*).mp | 6966647 |
| 8. | (colorectal or bowel or breast or mammar* or cervix or cervic*).mp | 2167242 |
| 9. | 7 and 8 | 1460608 |
| 10. | Colorectal Neoplasm*.mp. or exp colorectal tumor/ | 505106 |
| 11. | colorectal cancer.mp. or exp colorectal cancer/ | 461369 |
| 12. | exp breast tumor/ or exp breast carcinoma/ or breast neoplasm*.mp.<br>or exp breast cancer/ | 721902 |
| 13. | exp uterine cervix tumor/ or exp uterine cervix cancer/ or exp uterine cervix<br>carcinoma/ or uterine cervical neoplasm*.mp. | 150879 |
| 14. | 9 or 10 or 12 or 13 | 1609625 |
| 15. | ((((fecal or faecal) and test) or occult blood test or ((fecal or faecal) and<br>immunochemical test*) or (colonoscop* or endoscop* or sigmoid*) or<br>mammogra* or ((human and (papillomavirus or papilloma virus)) or<br>HPV)).mp | 899006 |
| 16. | exp Occult Blood/ | 11512 |
| 17. | exp Colonoscopy/ | 115984 |
| 18. | exp Endoscopy/ | 860222 |
| 19. | exp Sigmoidoscopy/ | 15151 |
| 20. | exp Mammography/ | 73066 |
| 21. | (screen* or early detect*).mp | 1997579 |
| 22. | exp Mass Screening/ | 343583 |
| 23. | "early detection of cancer".mp. or exp early cancer diagnosis/ | 19261 |
| 24. | 15 or 16 or 17 or 18 or 19 or 20 or 21 or 22 or 23 | 3203541 |
| 25. | 6 and 14 and 24 | 1945 |
| 26. | limit 25 to English language | 1912 |

| Web of Science Core Collection (Clarivate), 1900- |  |  |
| --- | --- | --- |
| # | Search terms | Results |
| 1. | ALL=(((intellectual or learning or neurodevelop* or development* or mental) AND (disabilit* or impair* or handicap or retard* or deficien* or disorder))) | 1,403,762 |
| 2. | ALL=((cancer* or neoplasm* or carcinoma* or tumor* or tumour* or oncolog*) AND (colorectal or bowel or breast or mammar* or cervix or cervic*)) | 1,310,307 |
| 3. | (((ALL=((fecal or faecal) and test)) OR ALL=("occult blood test")) OR ALL=((fecal or faecal) and "immunochemical test*")) OR ALL=(colonoscop* or endoscop* or sigmoid* or mammogra*) OR ALL=(papillomavirus or "papilloma virus" or HPV) | 659,001 |
| 4. | ALL=(screen* or "early detect*") | 1,562,020 |
| 5. | #4 AND #3 AND #2 AND #1 | 373 |
| 6. | #4 AND #3 AND #2 AND #1 and English (Languages) | 361 |

| PsycInfo (OVID), 1806- |  |  |
| --- | --- | --- |
| # | Search terms | Results |
| 1. | ((intellectual or learning or neurodevelop* or development* or mental) adj4 (disabilit* or impair* or handicap or retard* or deficient*)).mp. [mp=title, abstract, heading word, table of contents, key concepts, original title, tests & measures, mesh word] | 123456 |
| 2. | exp Intellectual Development Disorder/ | 51650 |
| 3. | intellectual disabilit*.mp. | 34531 |
| 4. | intellectual impairment.mp. | 1142 |
| 5. | exp Learning Disabilities/ or exp Learning Disorders/ or learning disorder*.mp. | 41749 |
| 6. | learning disabilit*.mp. [mp=title, abstract, heading word, table of contents, key concepts, original title, tests & measures, mesh word] | 29309 |
| 7. | developmental disabilit*.mp. | 23352 |
| 8. | (neurodevelopmental disabilit* or neurodevelopmental disorder*).mp. [mp=title, abstract, heading word, table of contents, key concepts, original title, tests & measures, mesh word] | 11043 |
| 9. | exp Developmental Disabilities/ or developmental disorder*.mp. | 23643 |
| 10. | 1 or 2 or 3 or 4 or 5 or 6 or 7 or 8 or 9 | 165890 |
| 11. | (cancer* or neoplasm* or carcinoma* or tumor* or tumour* or oncolog*).mp. [mp=title, abstract, heading word, table of contents, key concepts, original title, tests & measures, mesh word] | 108524 |
| 12. | (colorectal or bowel or breast or mammar* or cervix or cervic*).mp.<br>[mp=title, abstract, heading word, table of contents, key concepts,<br>original title, tests & measures, mesh word] | 44603 |
| 13. | 11 and 12 | 25199 |
| 14. | exp Colorectal Neoplasms/ or Colorectal Neoplasm*.mp. | 2698 |
| 15. | colorectal cancer.mp. | 3347 |
| 16. | exp Breast Neoplasms/ | 12461 |
| 17. | 13 or 14 or 15 or 16 | 25199 |
| 18. | ((fecal or faecal) and test) or occult blood test or ((fecal or faecal)<br>and immunochemical test*) or (colonoscop* or endoscop* or<br>sigmoid*) or mammogra* or ((human and (papillomavirus or<br>papilloma virus)) or HPV)).mp. [mp=title, abstract, heading word,<br>table of contents, key concepts, original title, tests & measures,<br>mesh word] | 8966 |
| 19. | occult blood.mp. | 468 |
| 20. | exp Mammography/ | 1477 |
| 21. | (screen* or early detect*).mp. [mp=title, abstract, heading word,<br>table of contents, key concepts, original title, tests & measures,<br>mesh word] | 167217 |
| 22. | exp Cancer Screening/ or exp Screening/ or exp Screening Tests/ | 28347 |
| 23. | 18 or 19 or 20 or 21 or 22 | 173707 |
| 24. | 10 and 17 and 23 | 76 |
| 25. | limit 24 to English language | 74 |

## Appendix 2. Organisations included in the website search (websites and results)

| **Learning disabilities** |  |  |
| --- | --- | --- |
| **Organisation** | **Website** | **Results of searches** |
| Mencap | https://www.mencap.org.uk/ | 98 |
| IDS Tilda | https://idstilda.tcd.ie/ | 20 |
| Inclusion Europe | https://www.inclusion-europe.eu/ | 23 |
| British Institute of Learning Disabilities | https://www.bild.org.uk/ | 5 |
| Foundation for People with Learning Disabilities | https://www.learningdisabilities.org.uk/ | 5 |

| **Bowel cancer** |  |  |
| --- | --- | --- |
| **Organisation** | **Website** | **Results of searches** |
| Bowel Cancer UK | https://www.bowelcanceruk.org.uk/ | 7 |
| Bowel Research UK | https://bowelresearchuk.org/ | 14 |
| Fight Colorectal Cancer | https://fightcolorectalcancer.org/ | 0 |
| Colorectal Cancer Alliance | https://colorectalcancer.org/ | 32 |

| **Breast cancer** |  |  |
| --- | --- | --- |
| **Organisation** | **Website** | **Results of searches** |
| Breast Cancer Now | https://breastcancernow.org | 31 |
| Breast Cancer Research Foundation | https://www.bcrf.org/ | 1 |
| Prevent breast cancer | https://preventbreastcancer.org.uk/ | 0 |
| Susan G Komen | https://www.komen.org/ | 58 |
| Pink Ribbon Foundation | https://www.pinkribbonfoundation.org.uk/ | 4 |
| National Breast Cancer Foundation | https://www.nationalbreastcancer.org/ | 3 |

| **Cervical cancer** |  |  |
| --- | --- | --- |
| **Organisation** | **Website** | **Results of searches** |
| The Eve Appeal | https://eveappeal.org.uk/ | 10 |
| British Gynaecological Cancer Society | https://www.bgcs.org.uk/ | 0 |

| **General cancer** |  |  |
| --- | --- | --- |
| **Organisation** | **Website** | **Results of searches** |
| Cancer Research UK | https://www.cancerresearchuk.org/ | 518 |
| International Agency for Research on Cancer | https://www.iarc.who.int/ | 13 |

## Appendix 3. Studies excluded at full text assessment

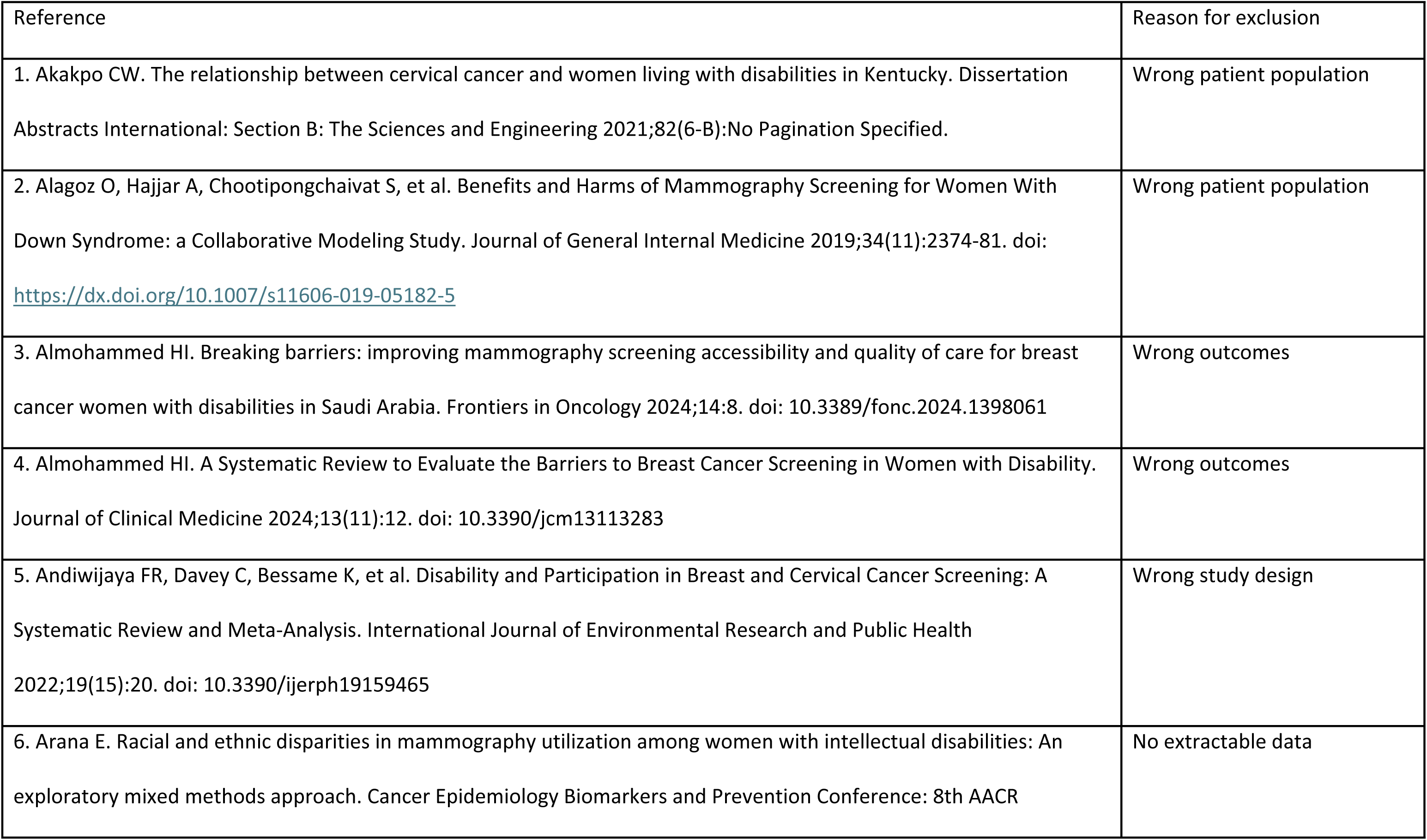
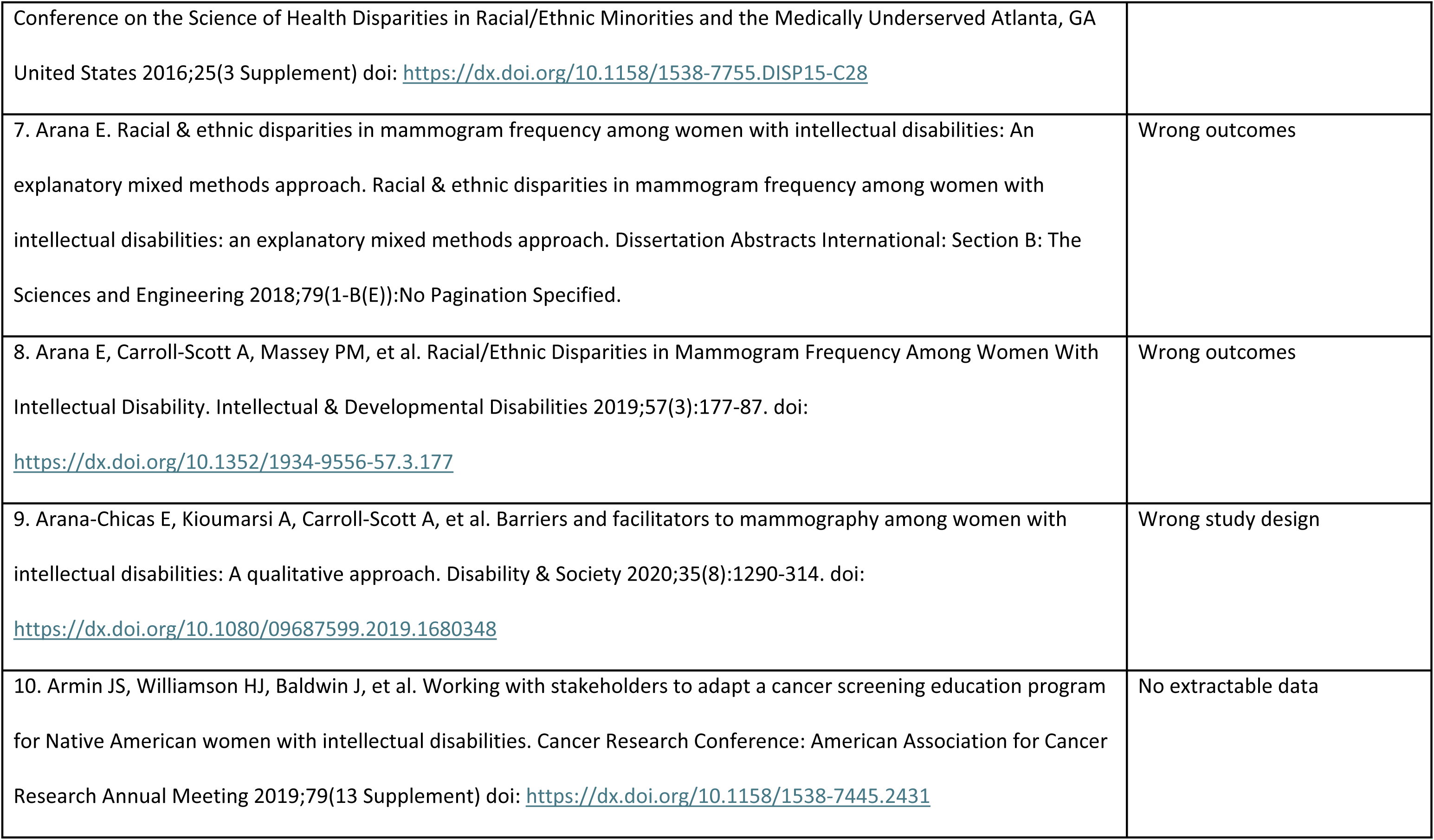
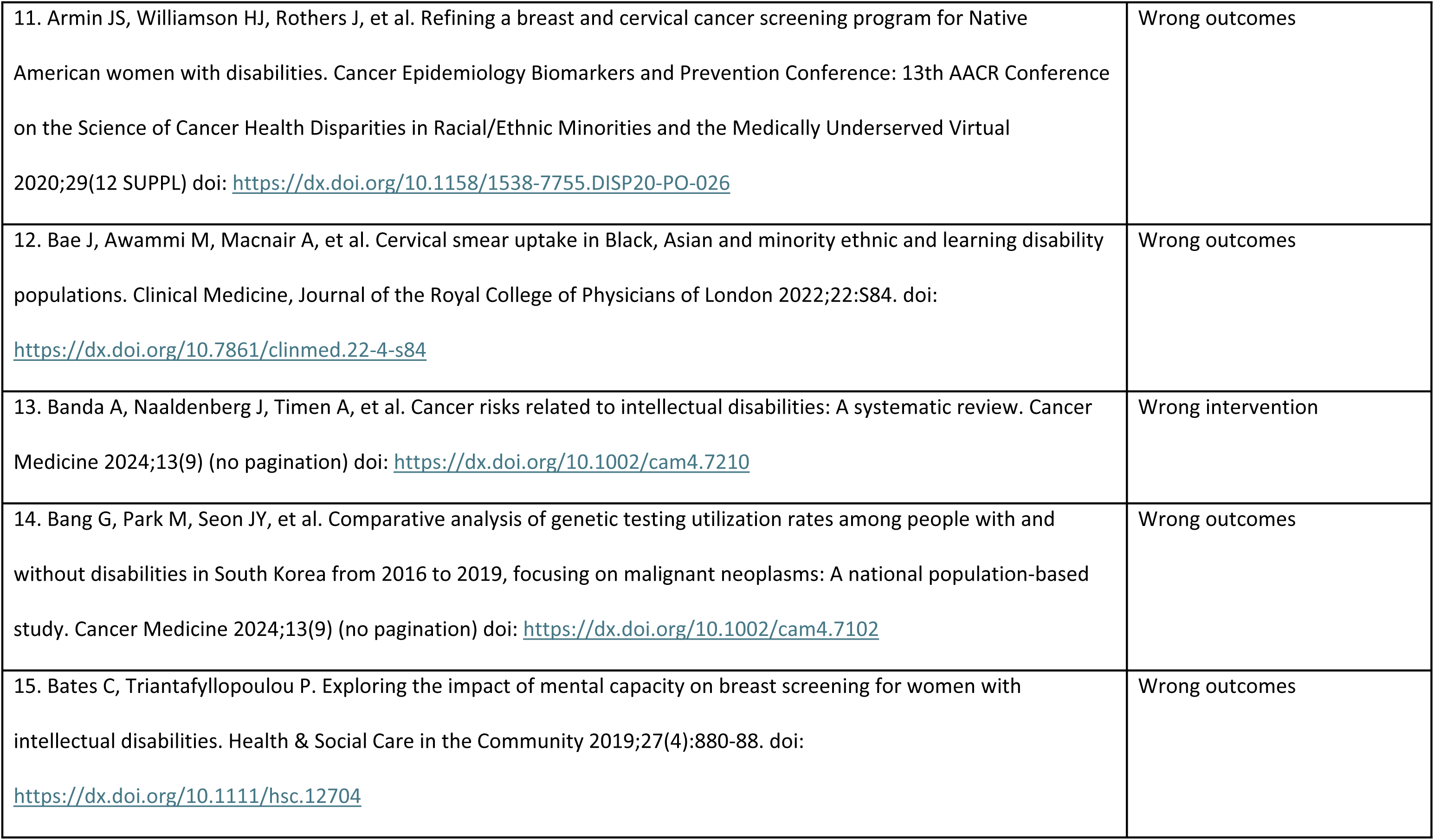
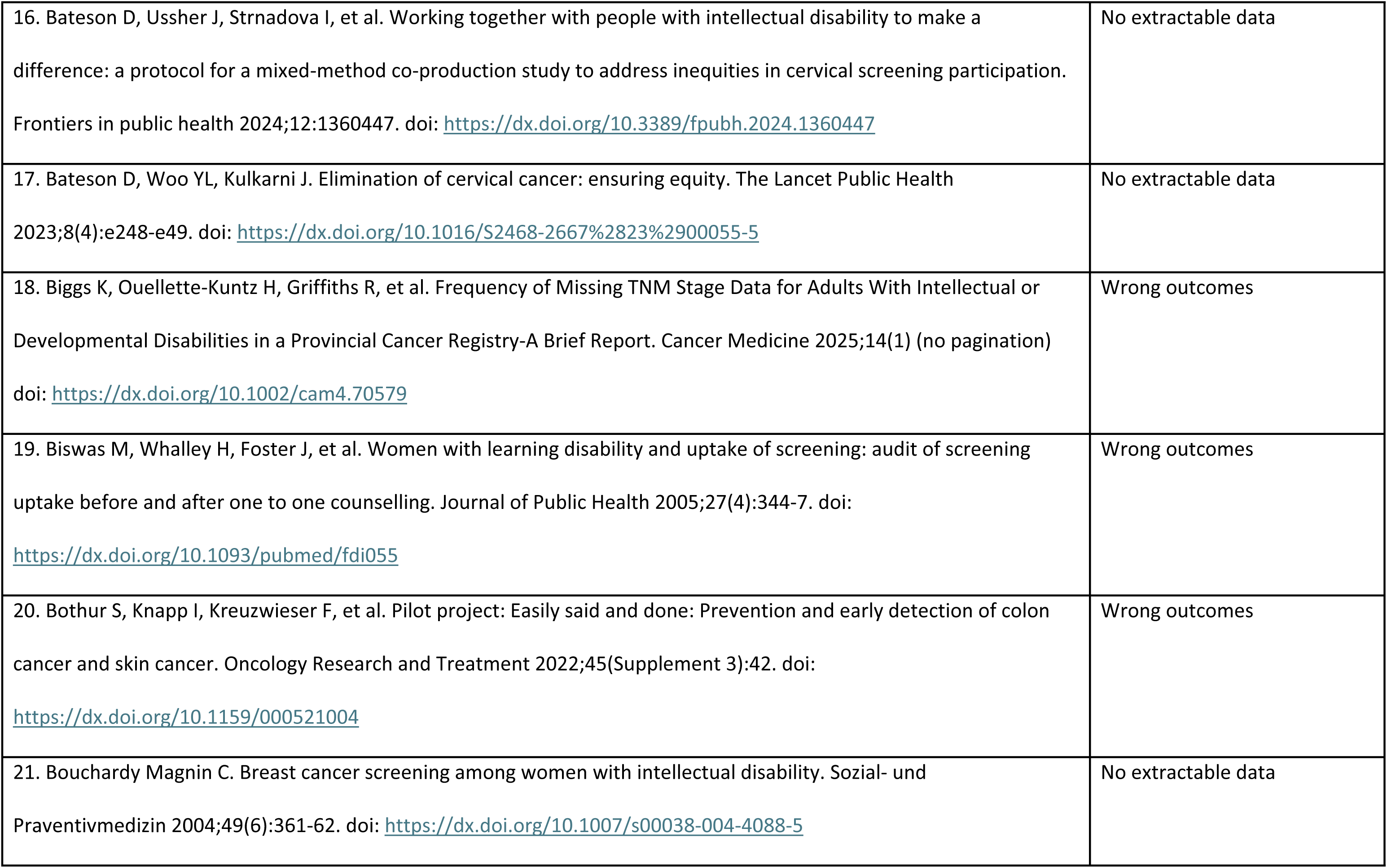
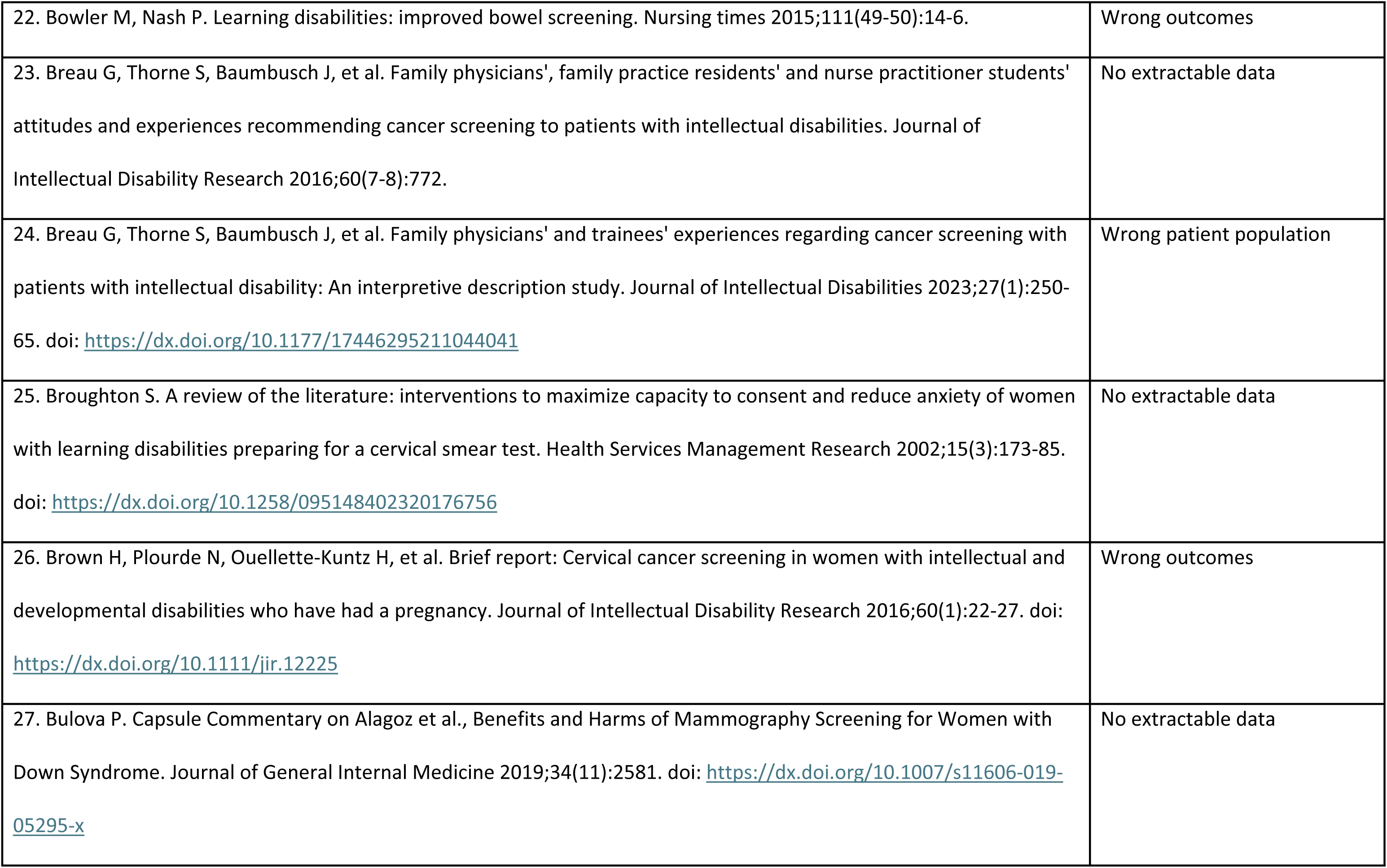
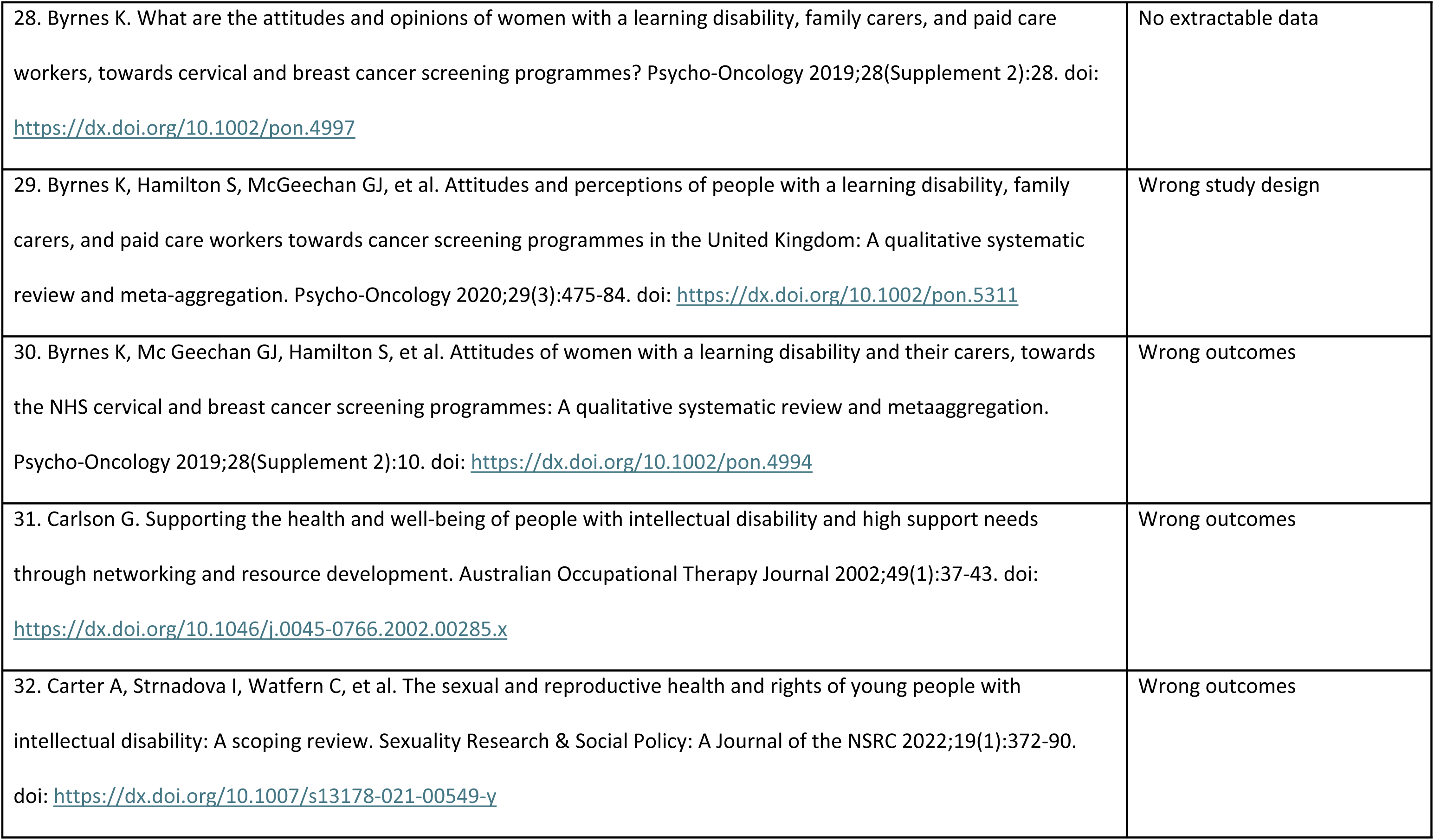
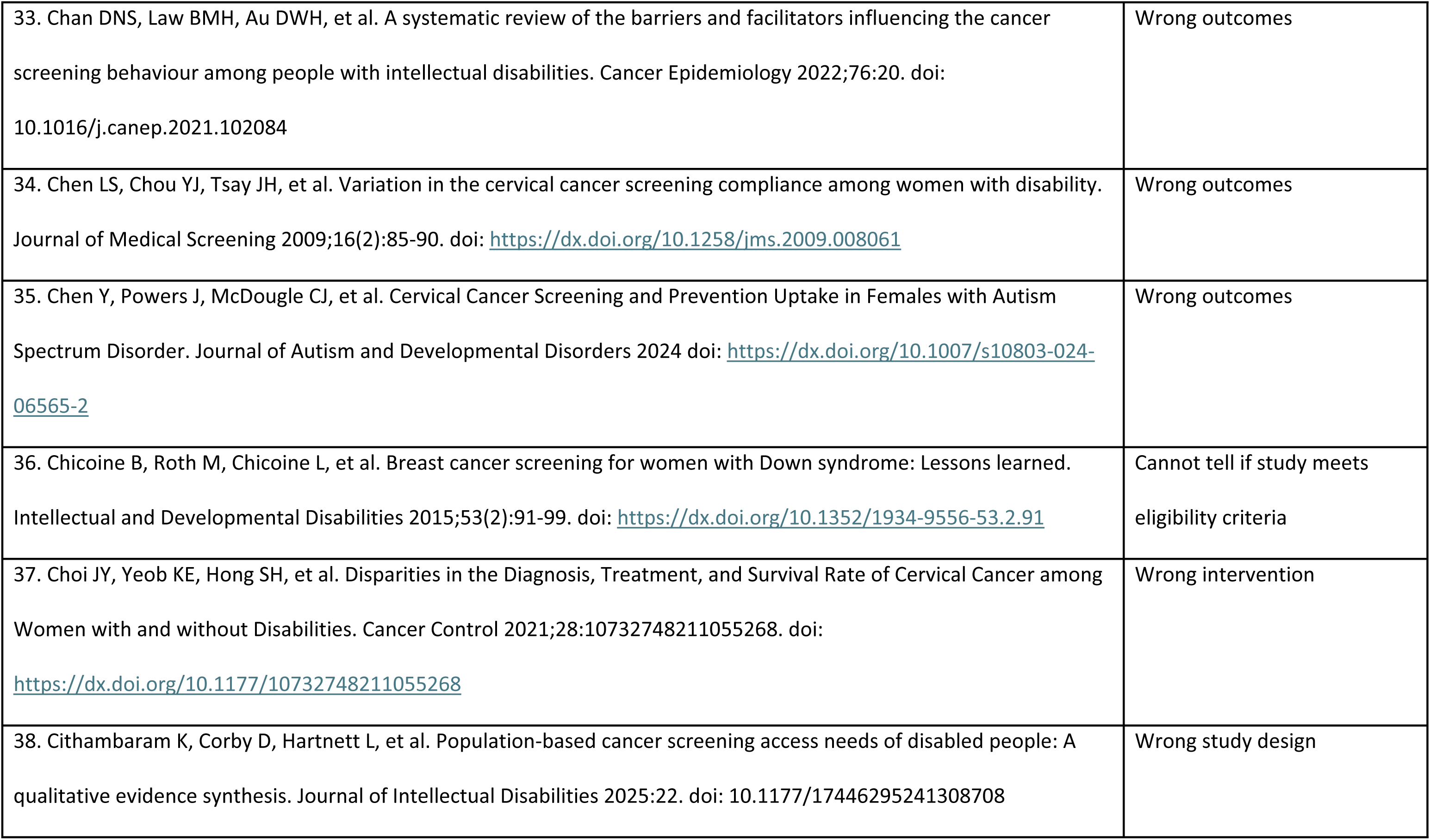
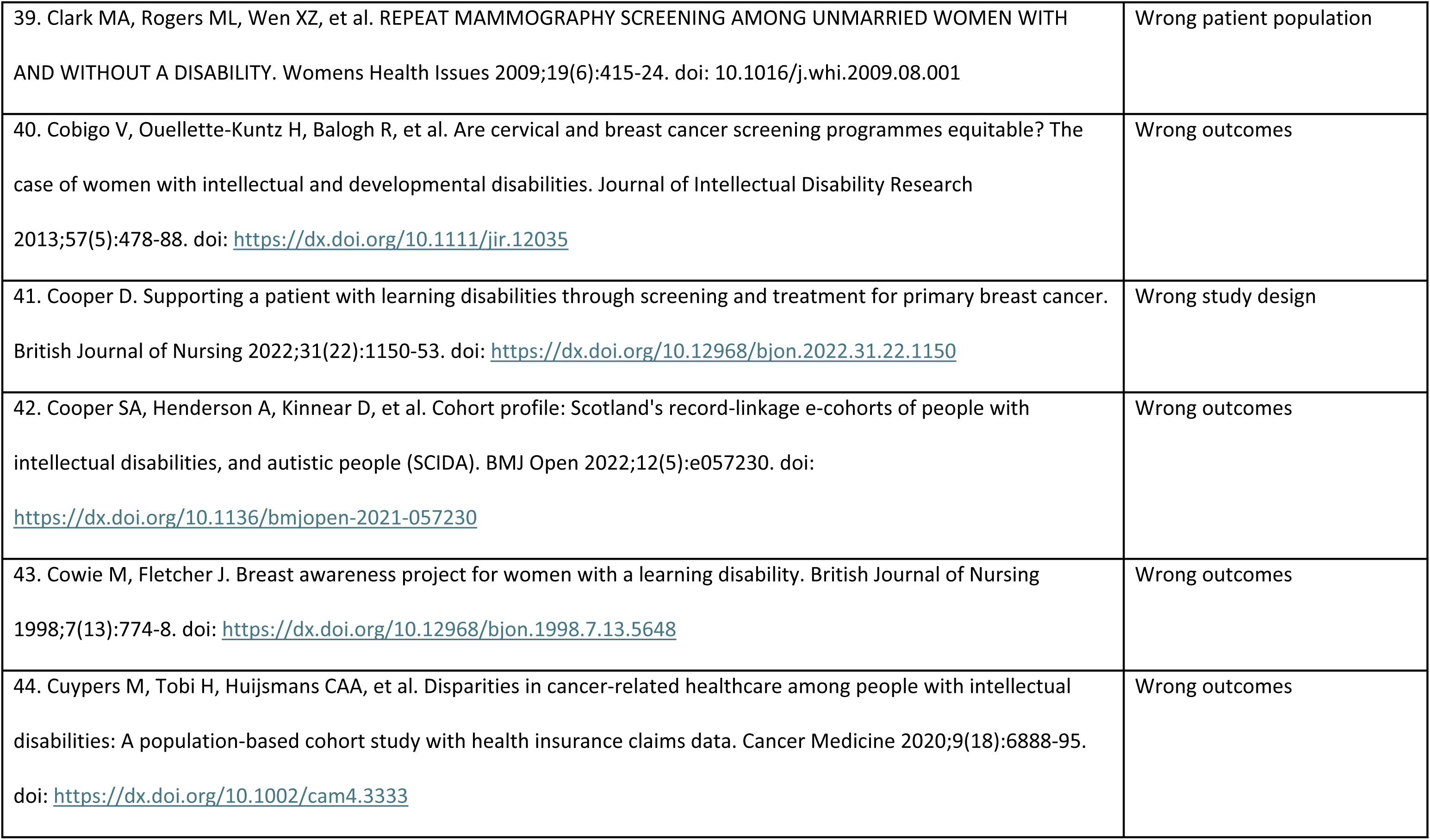
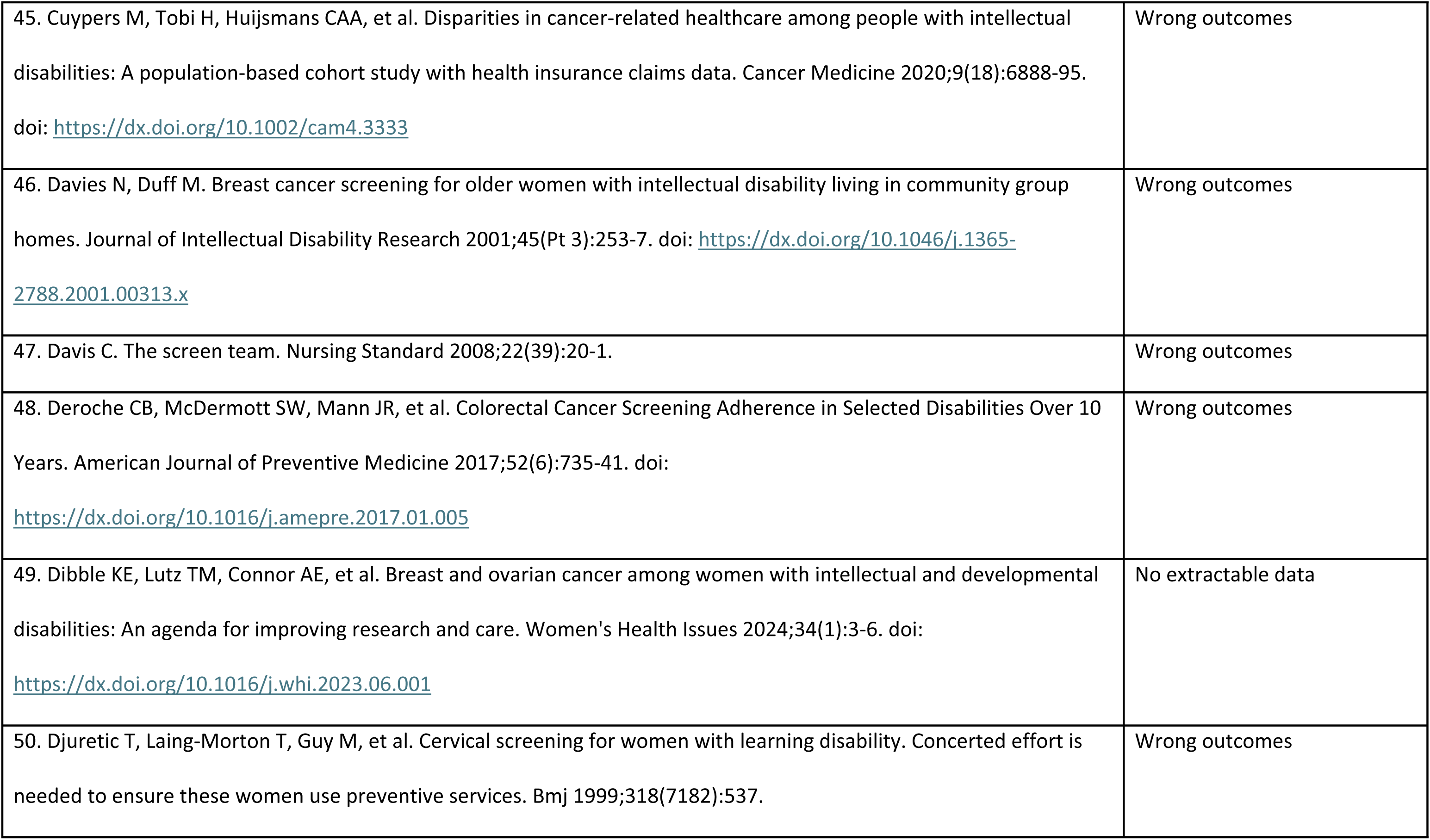
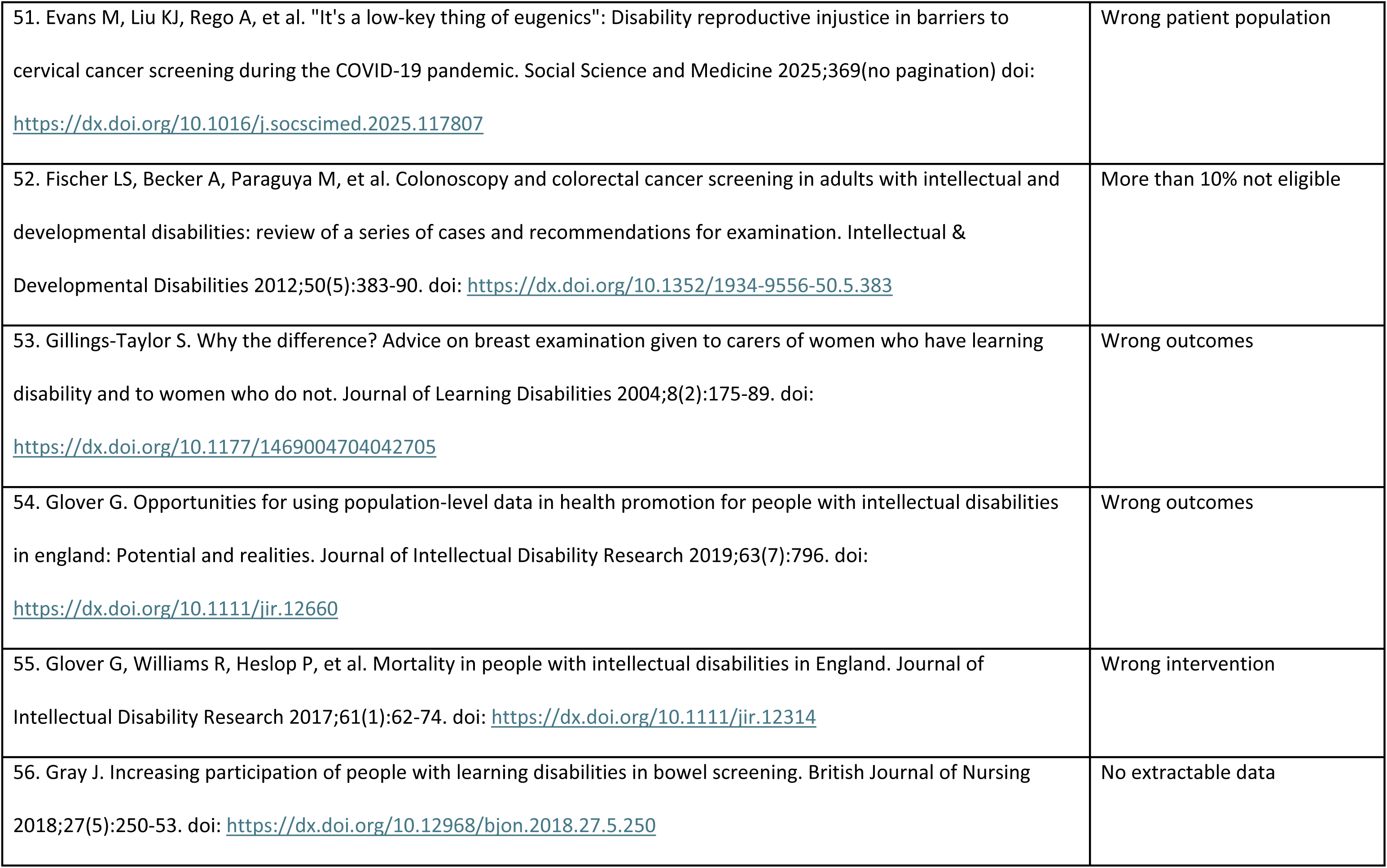
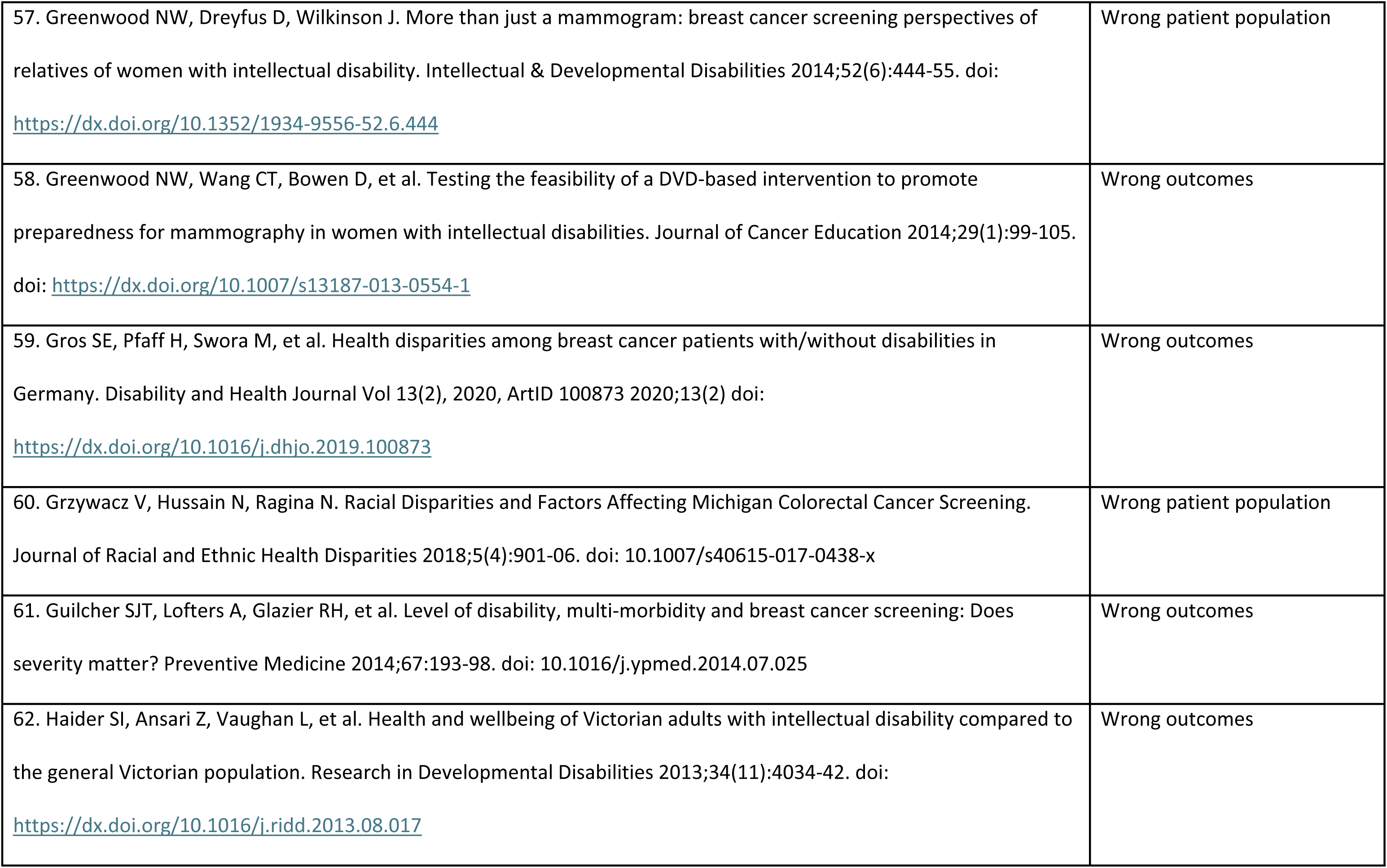
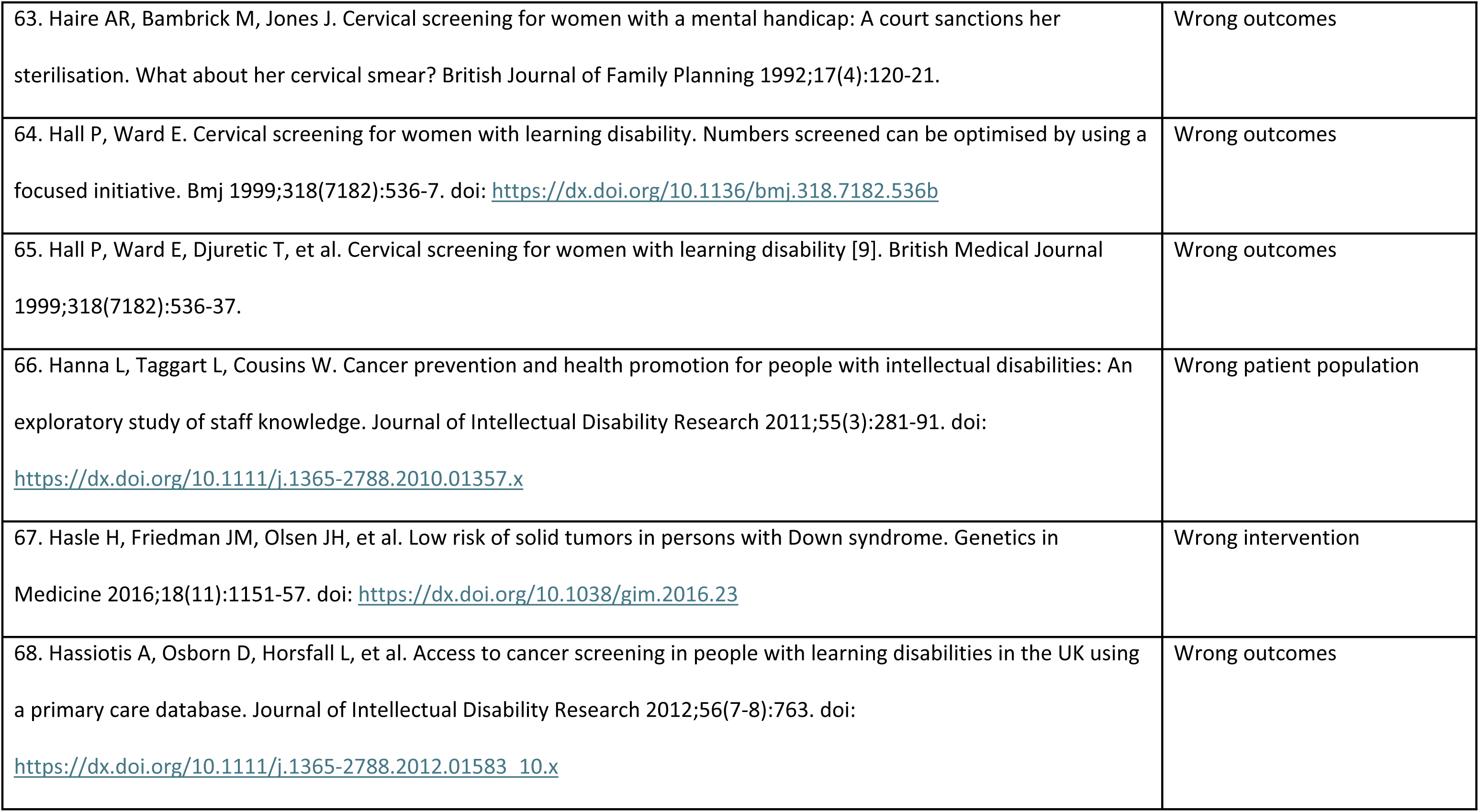
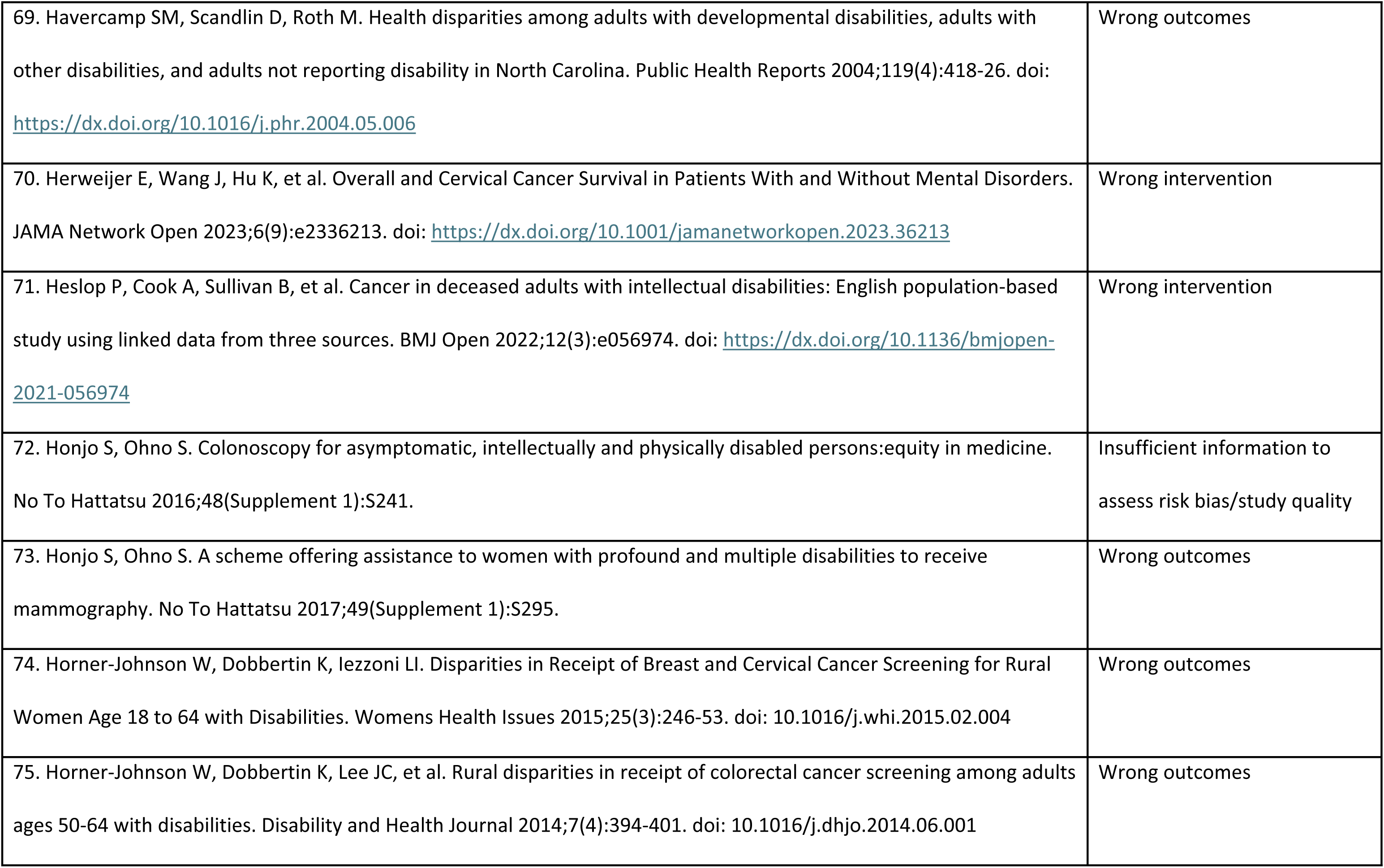
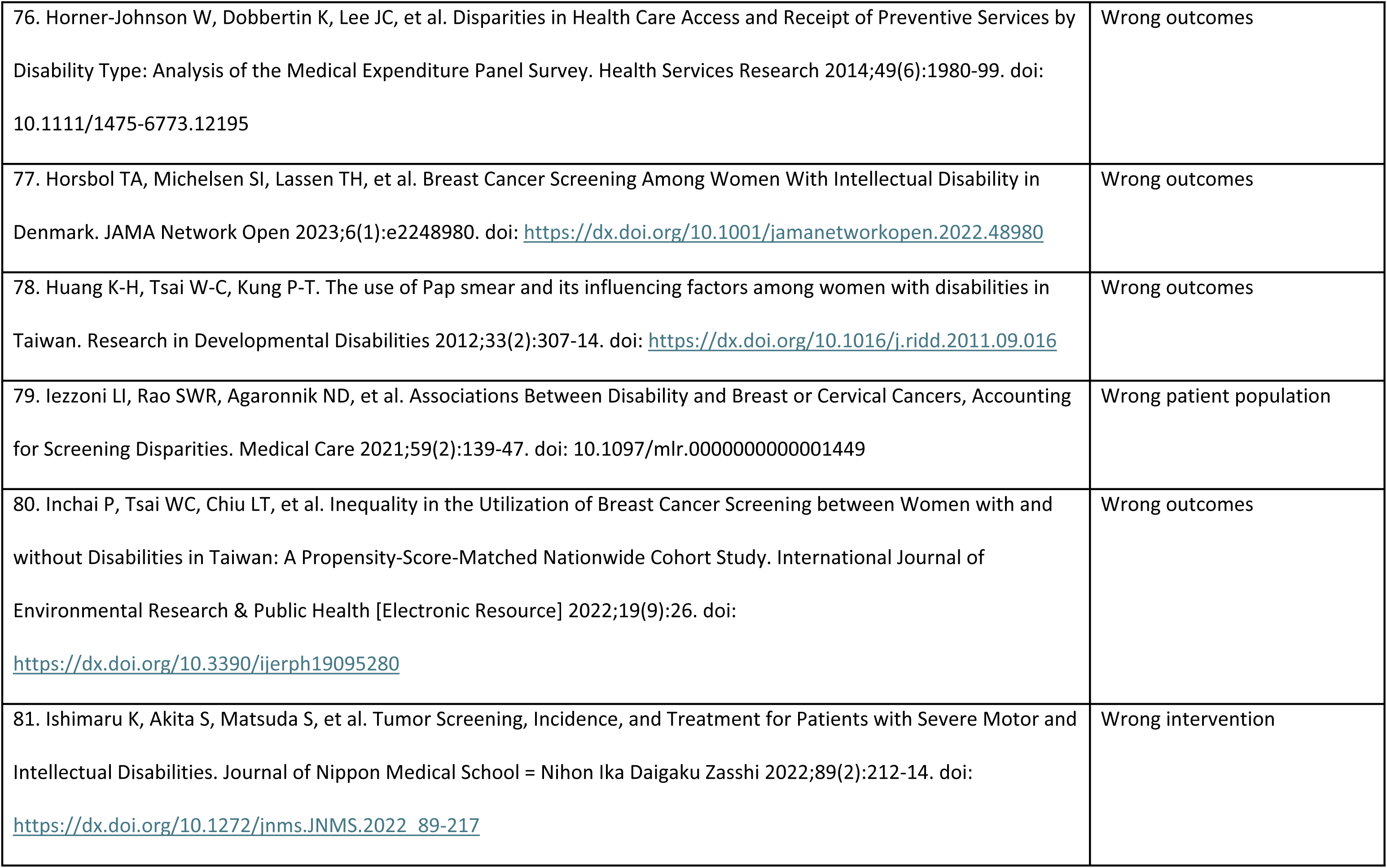
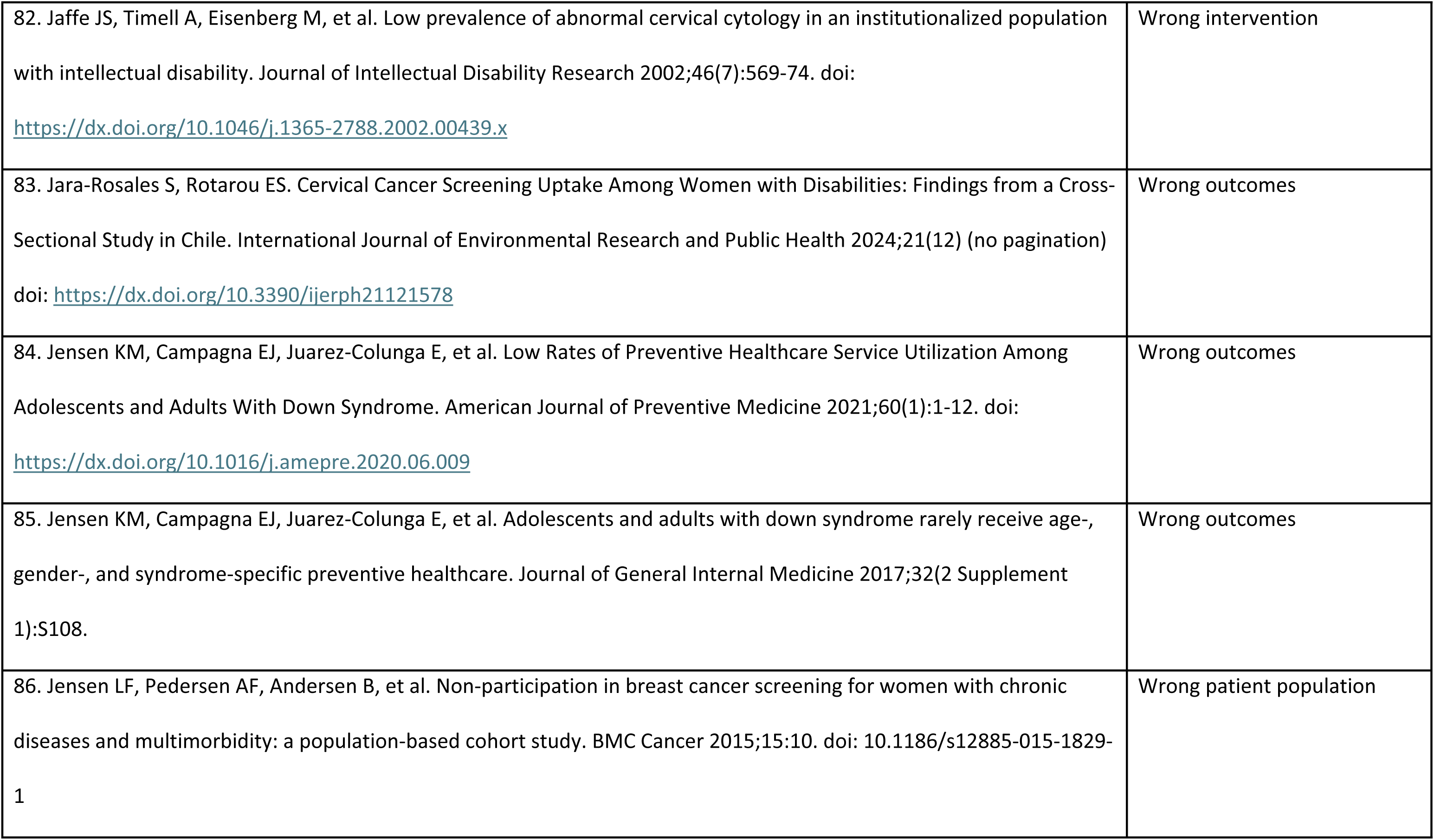
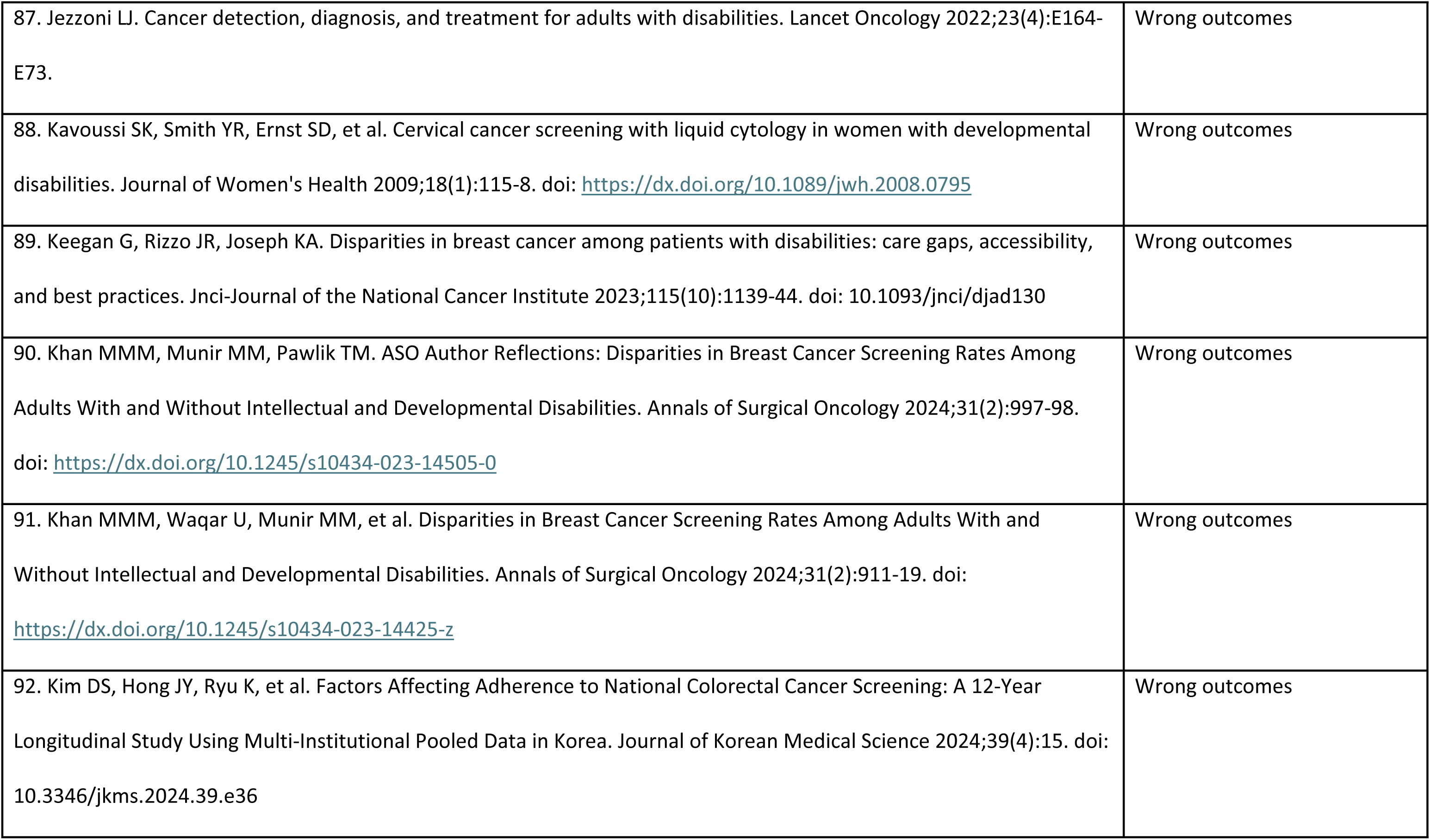
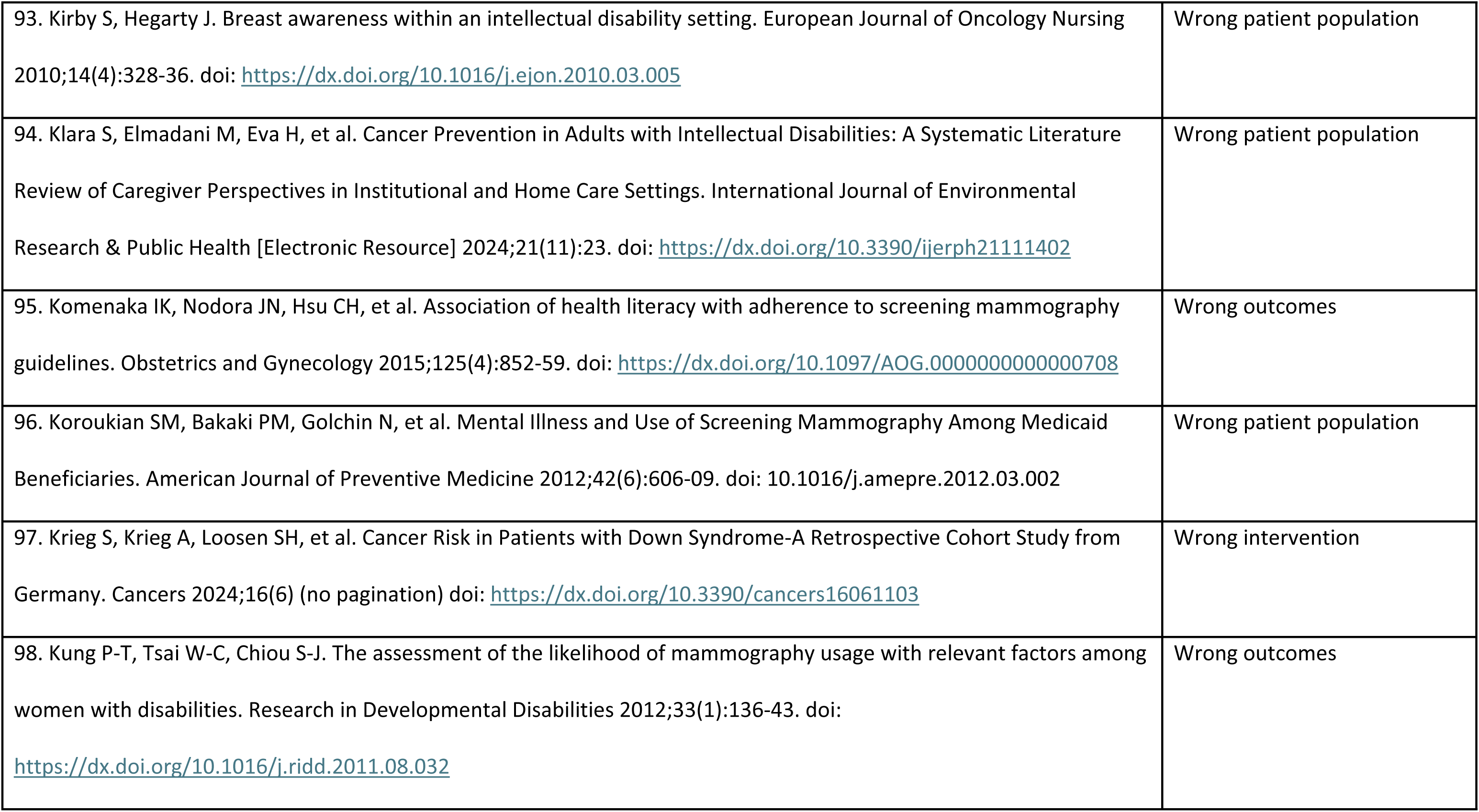
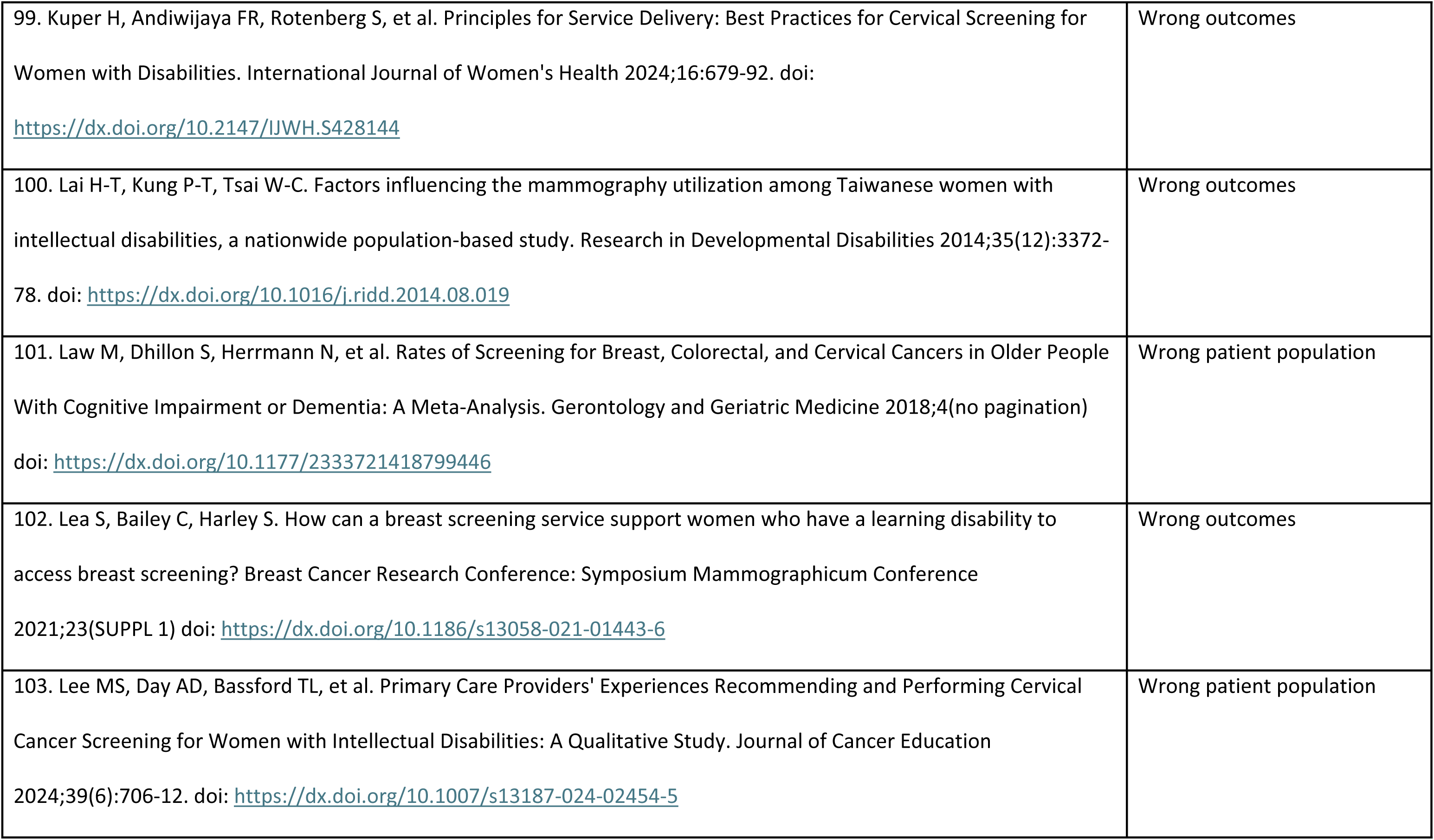
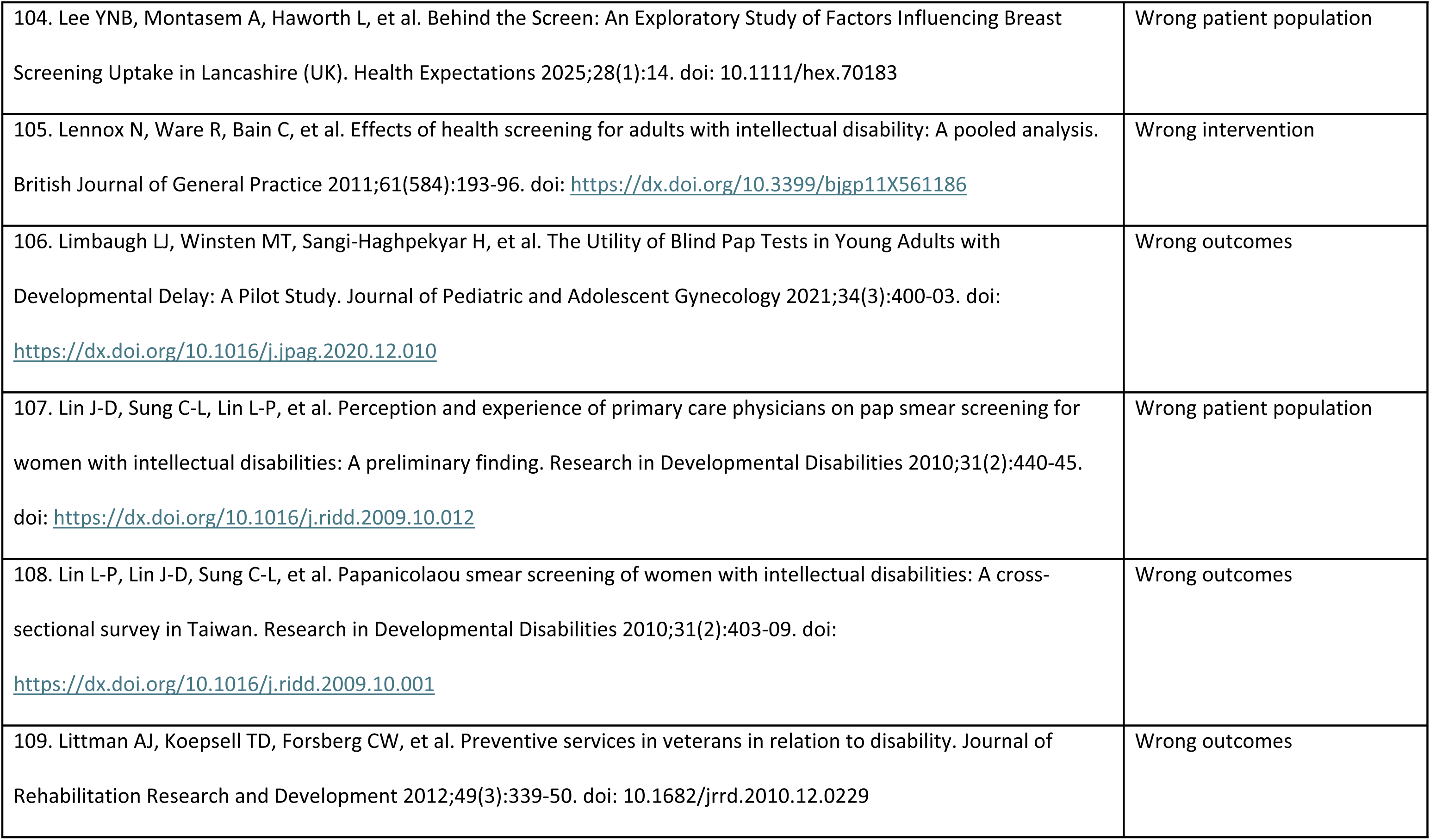
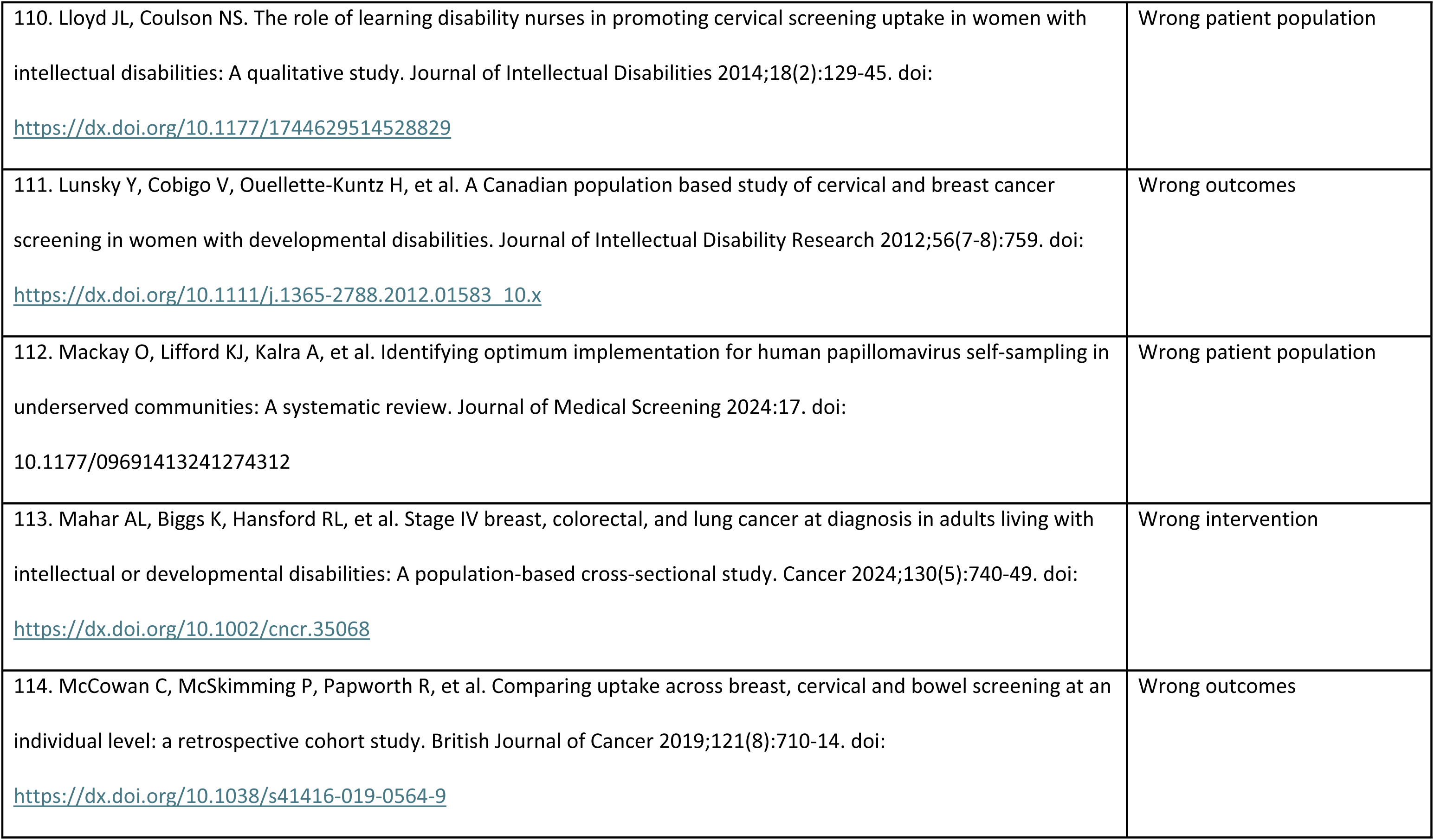
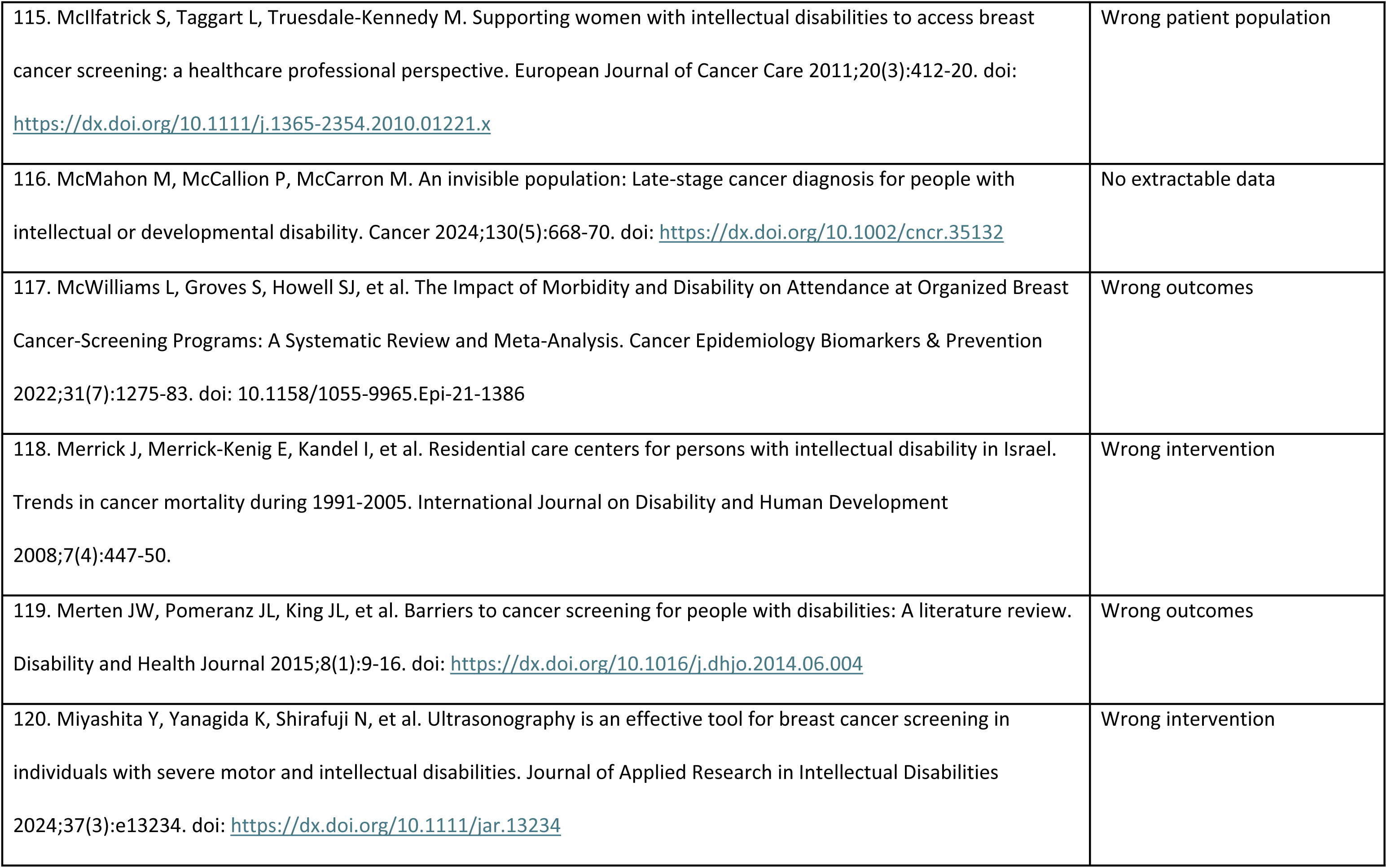
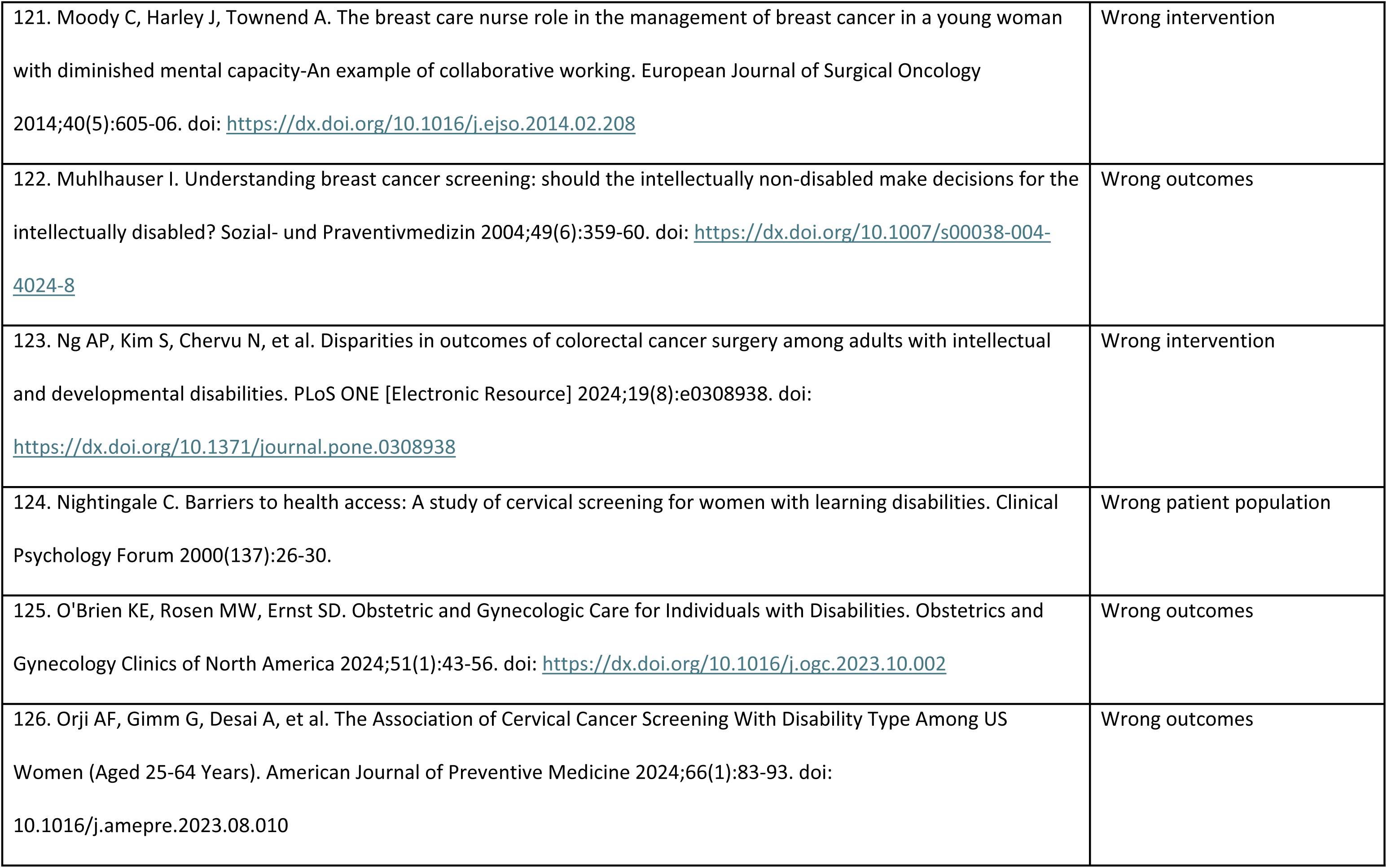
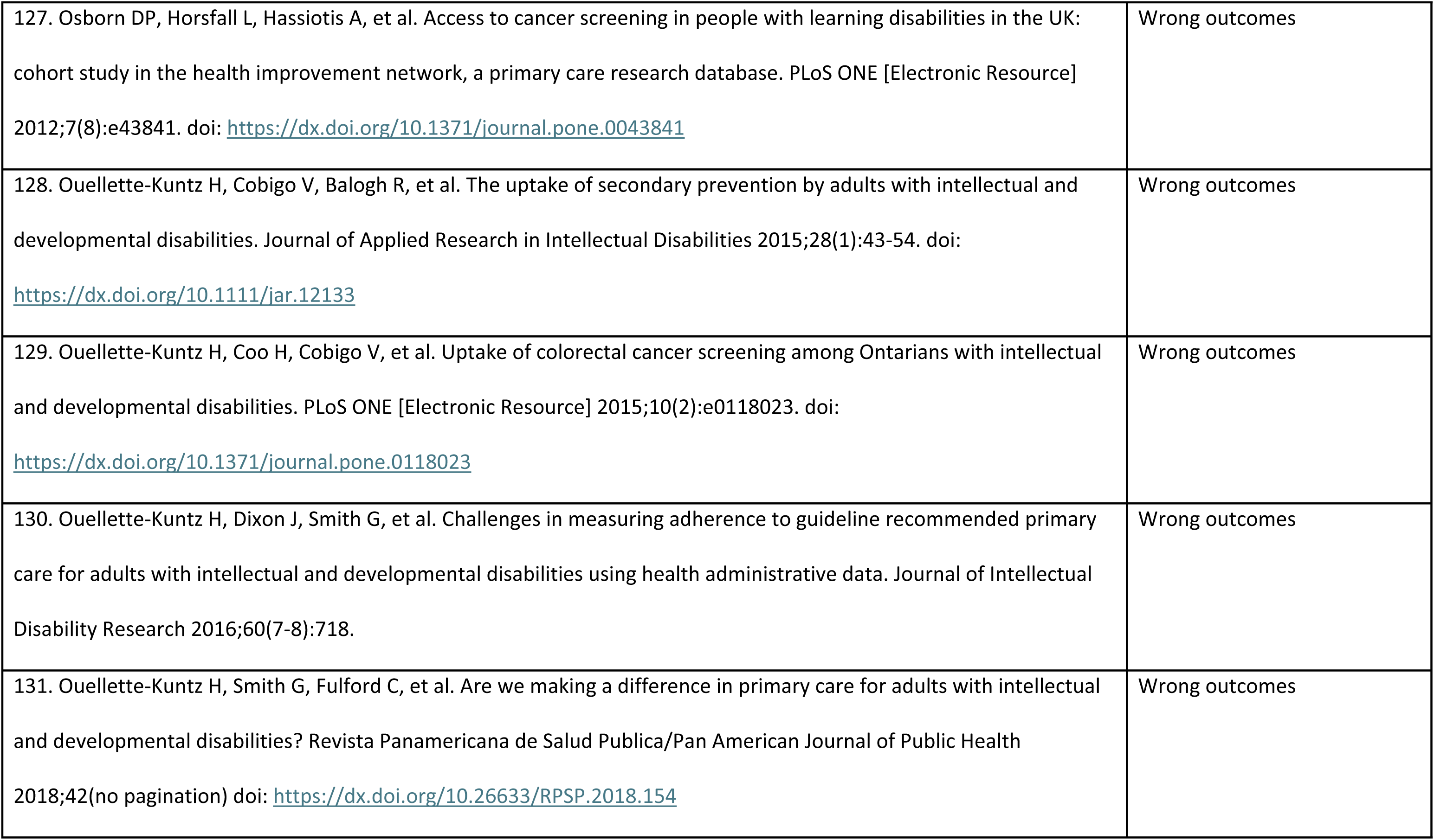
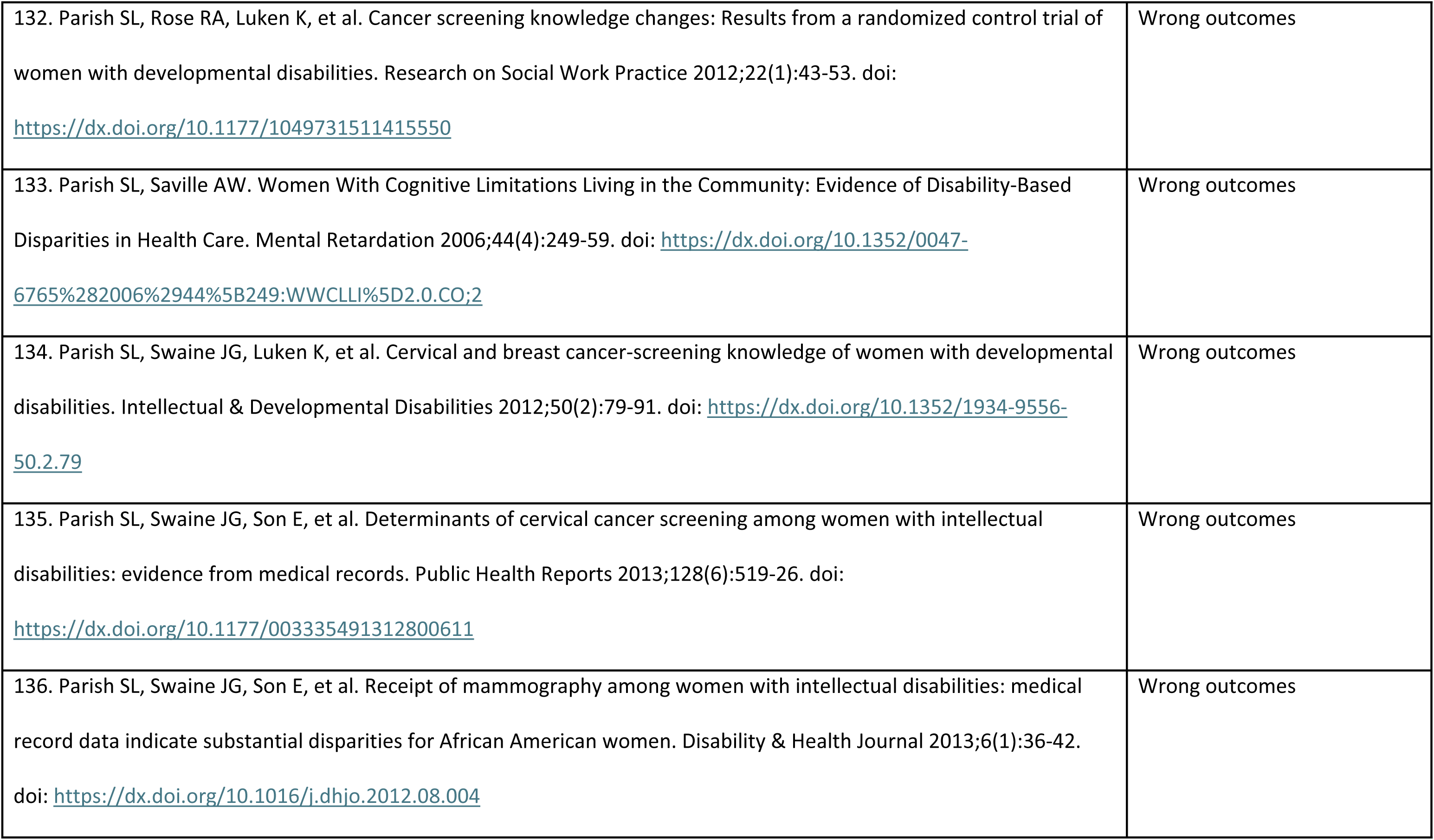
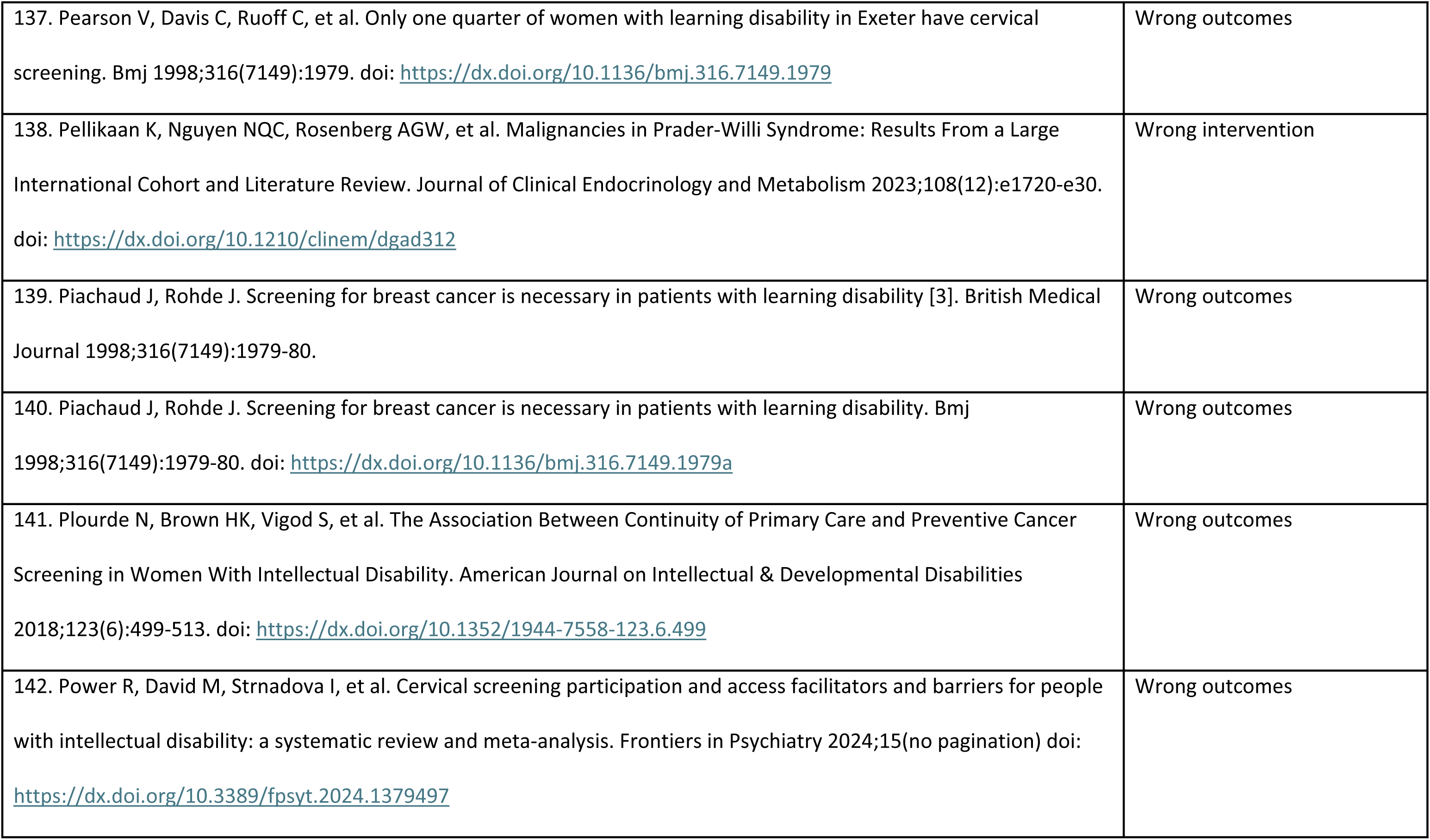
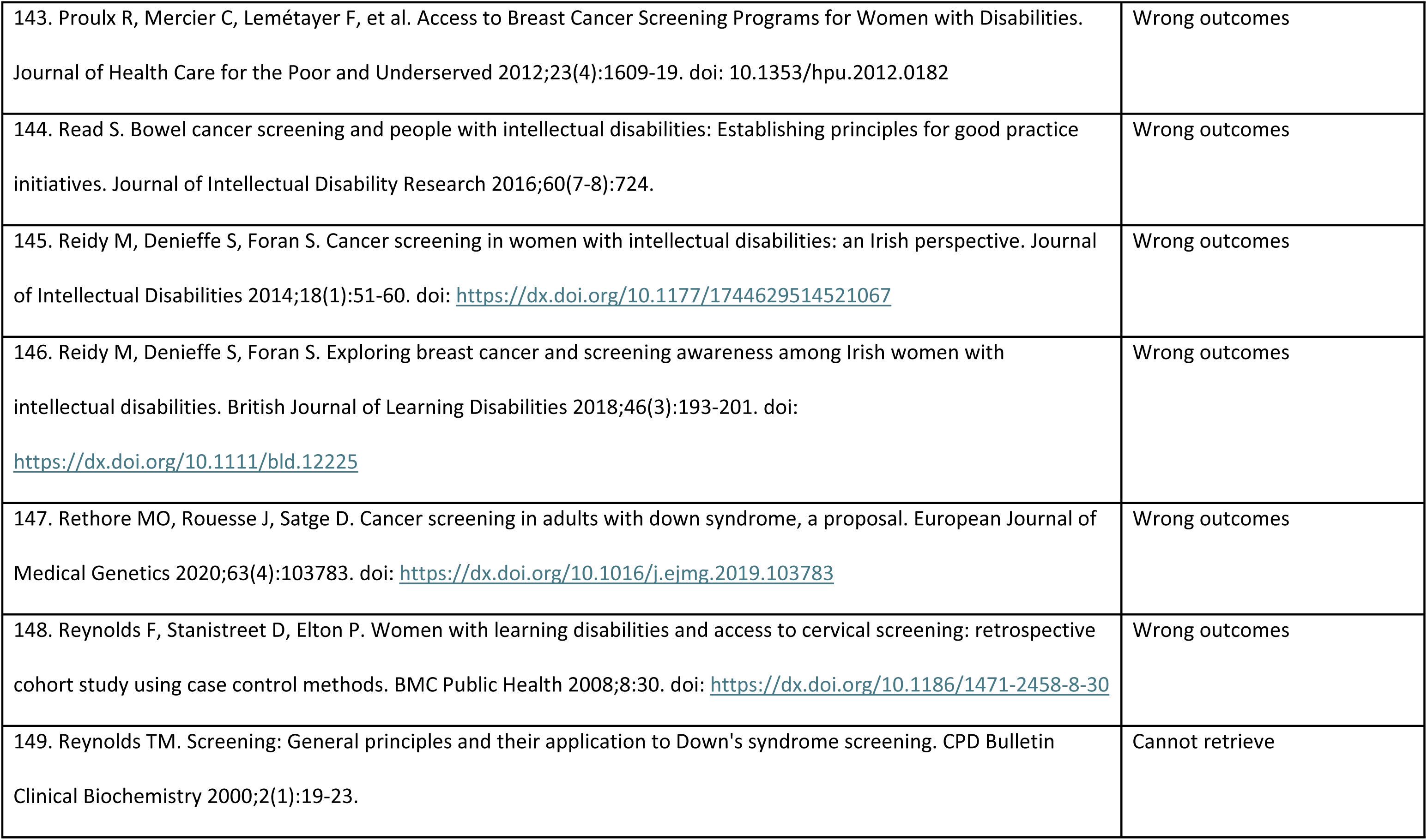
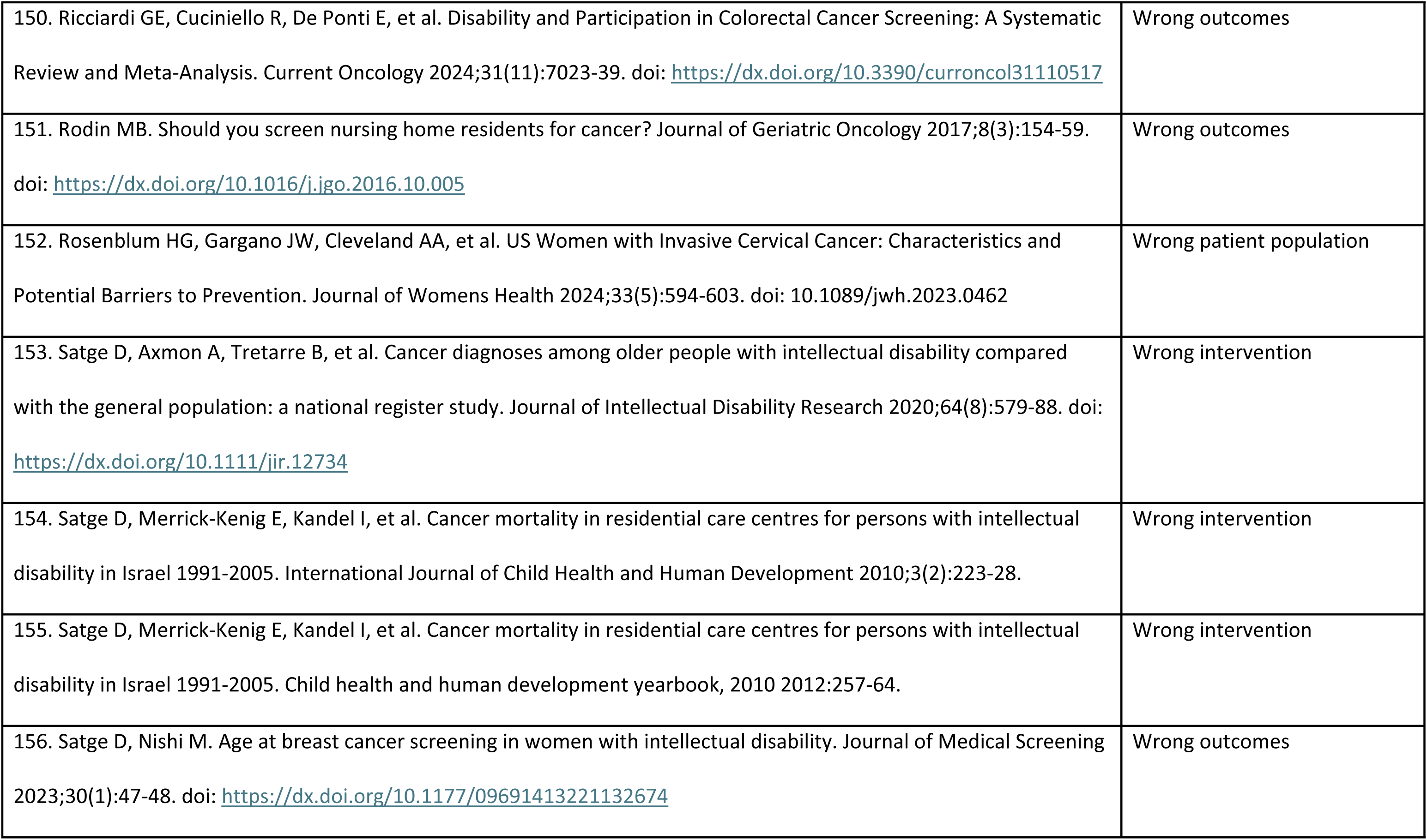
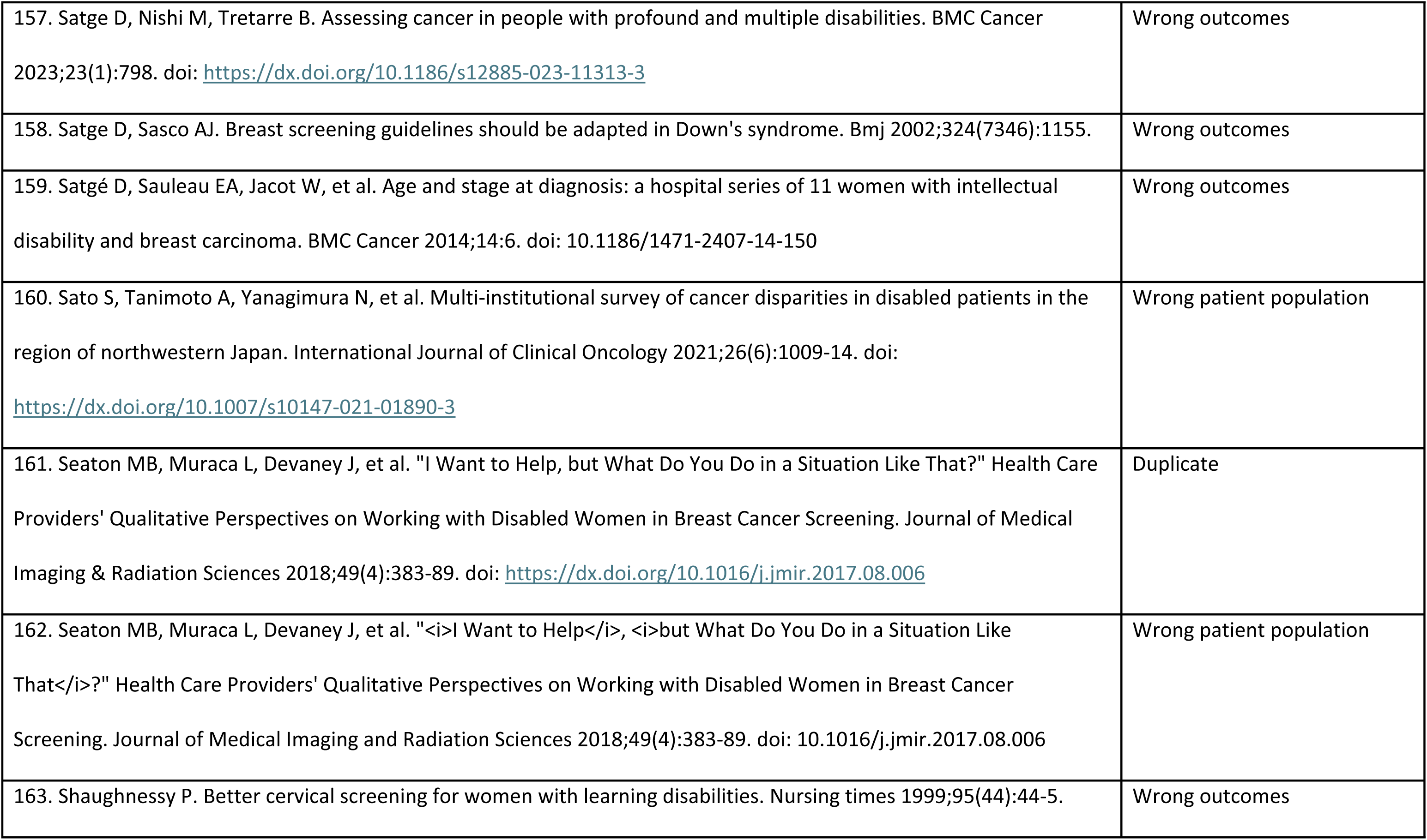
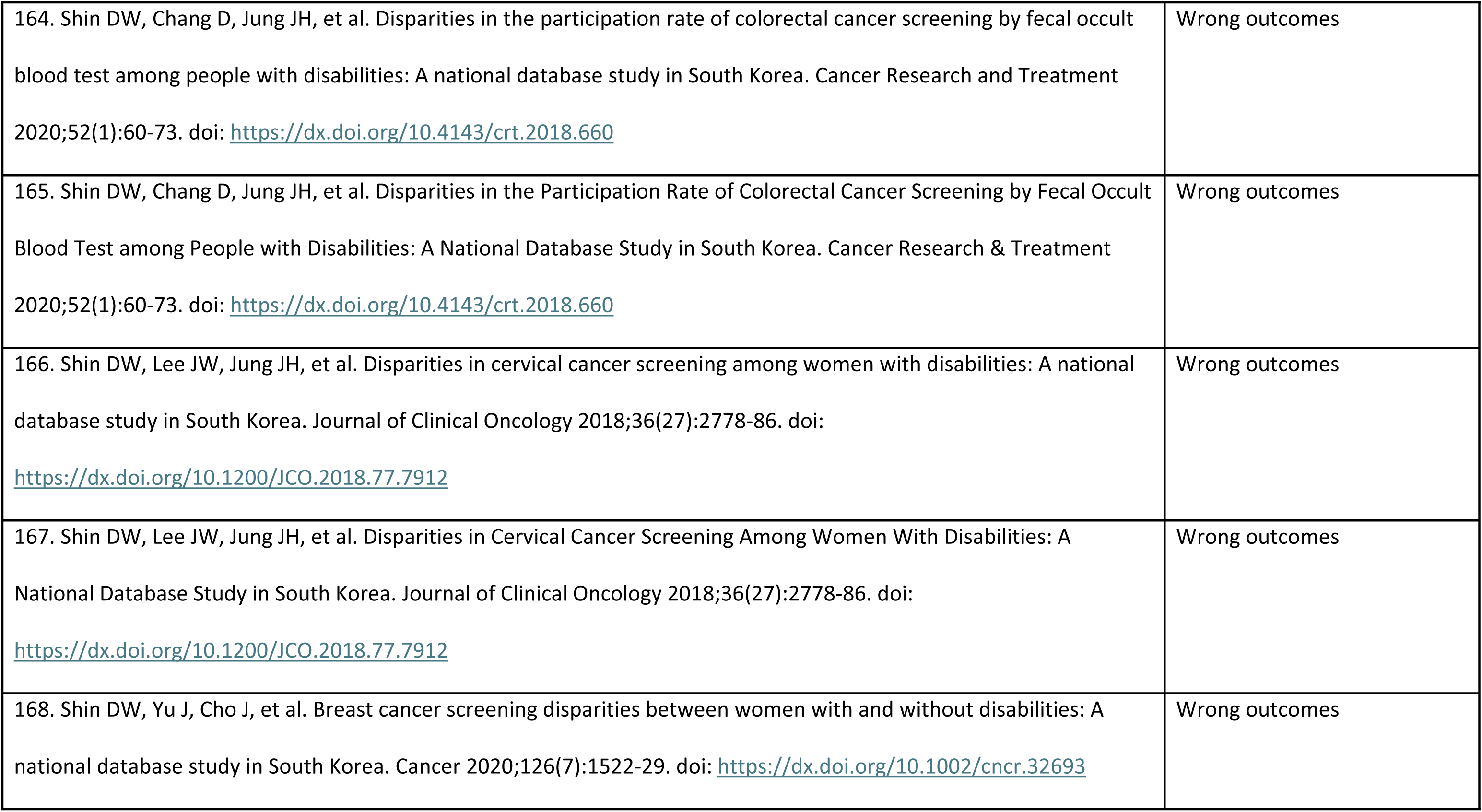
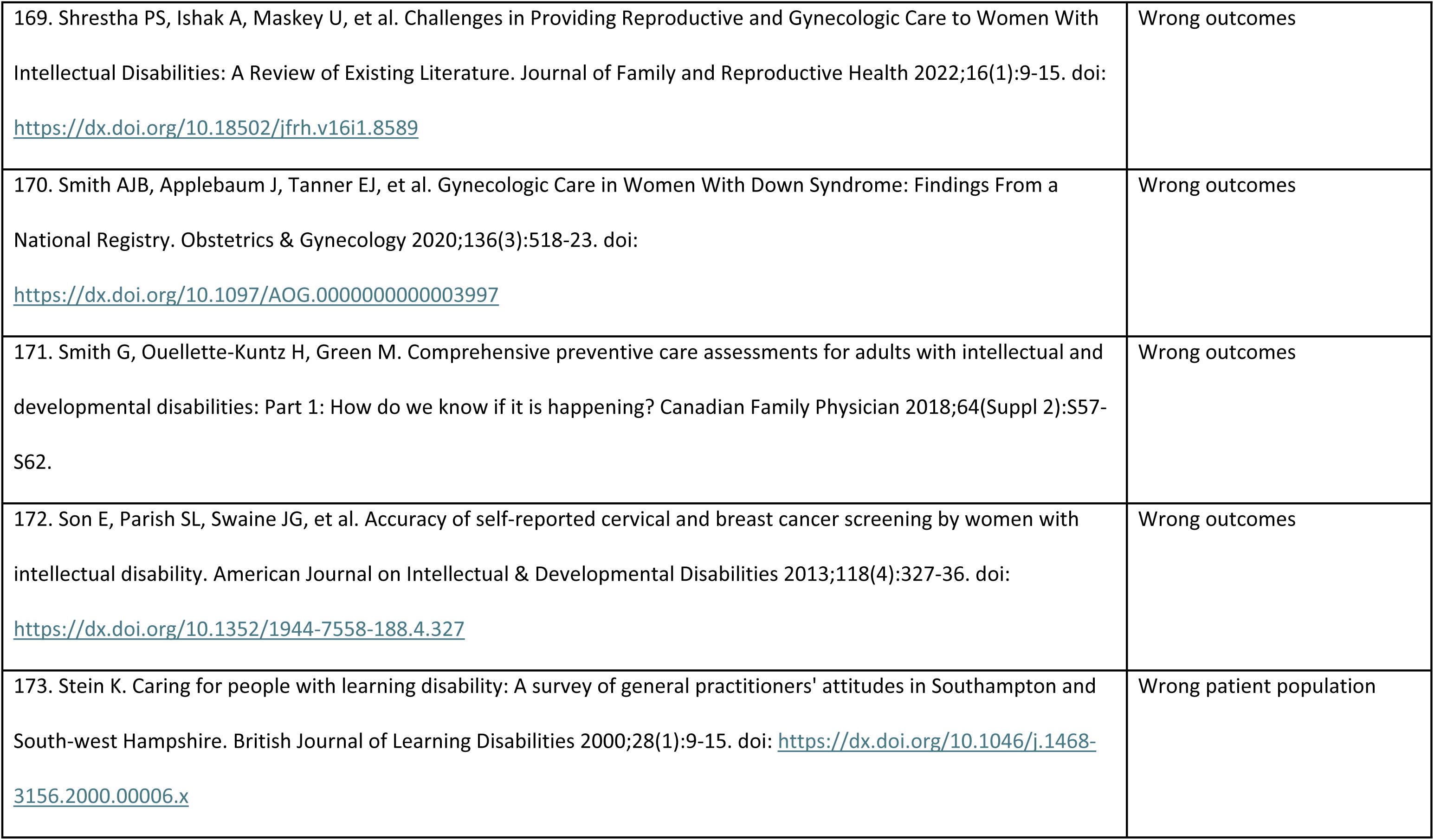
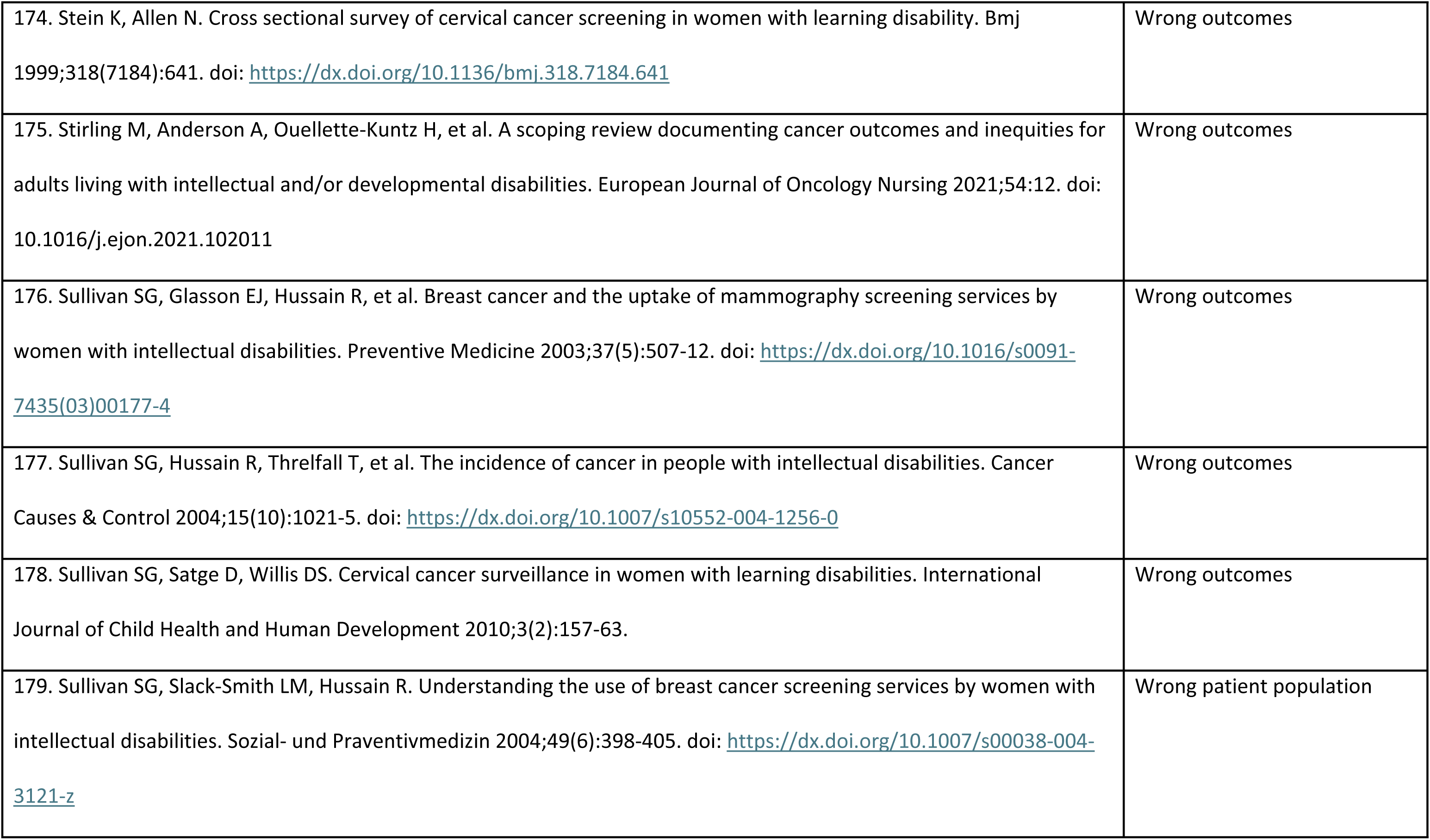
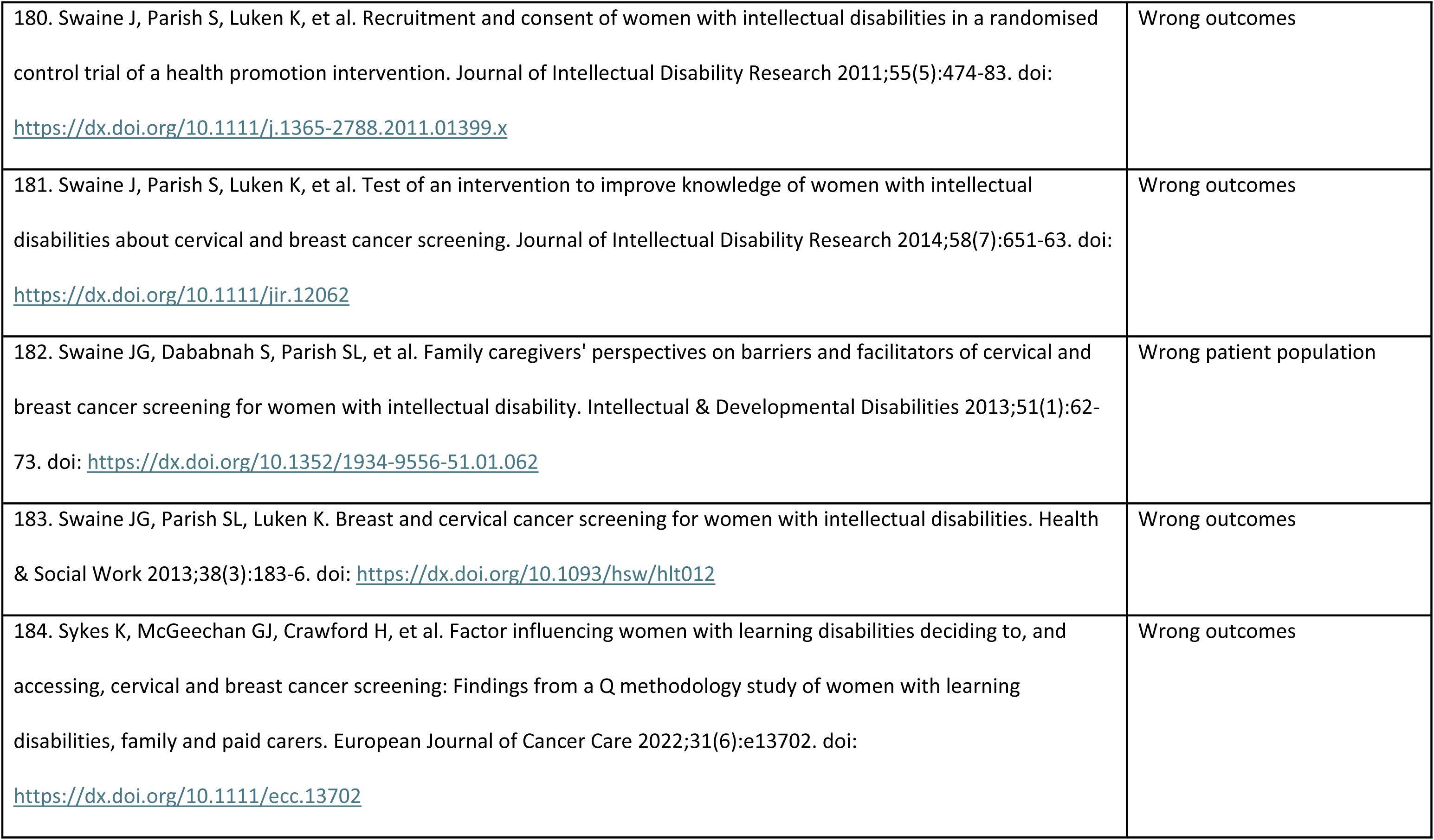
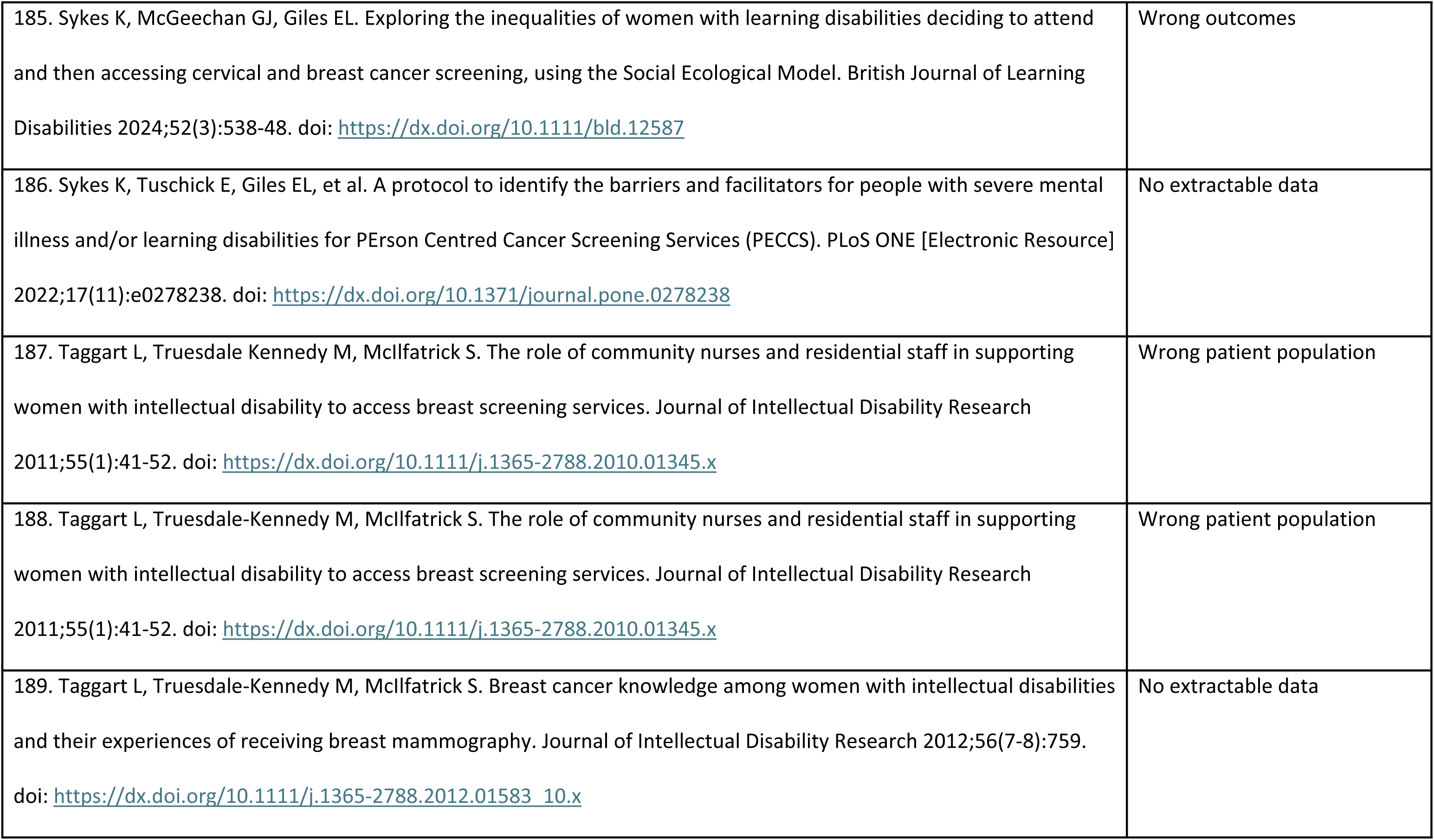
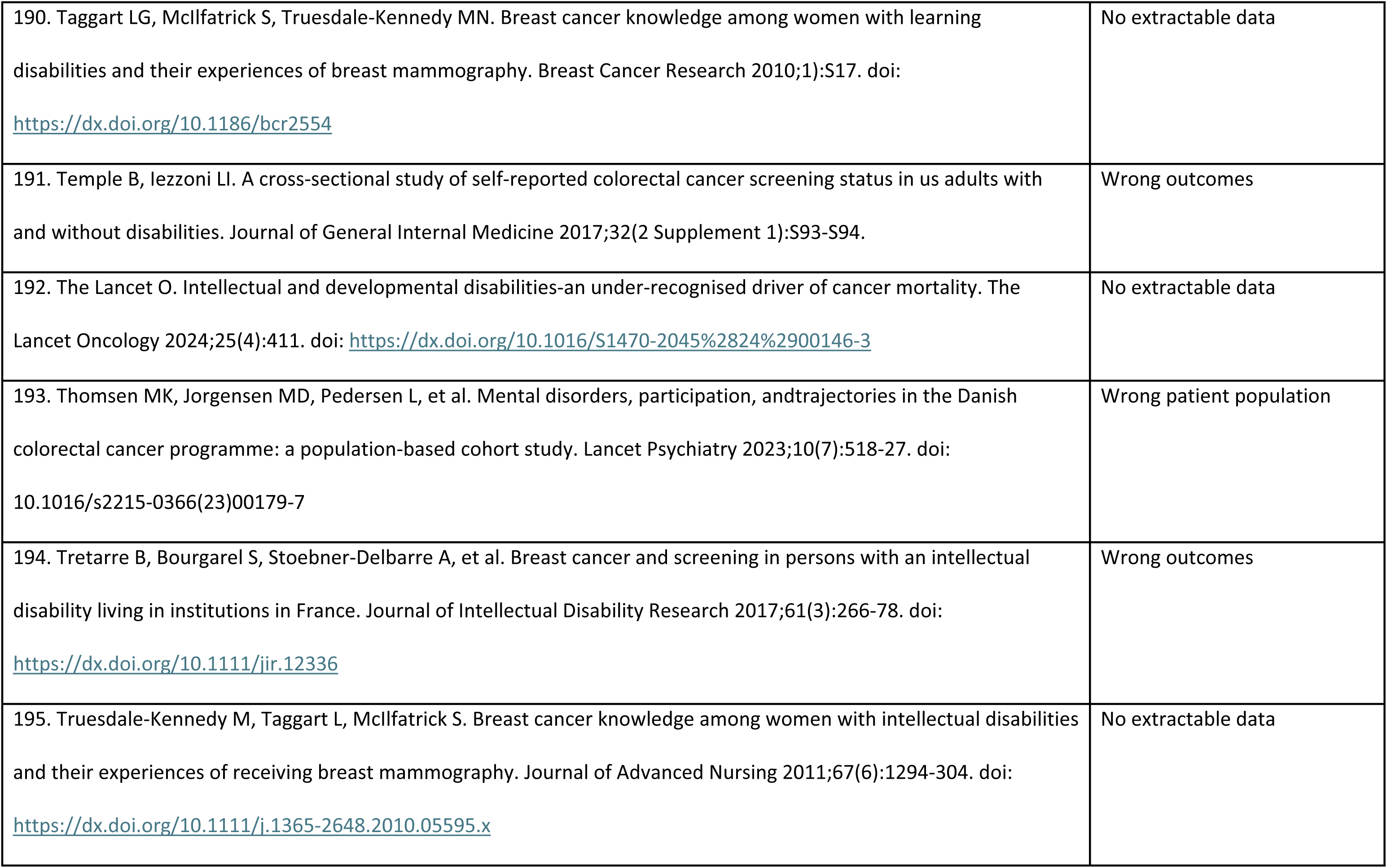
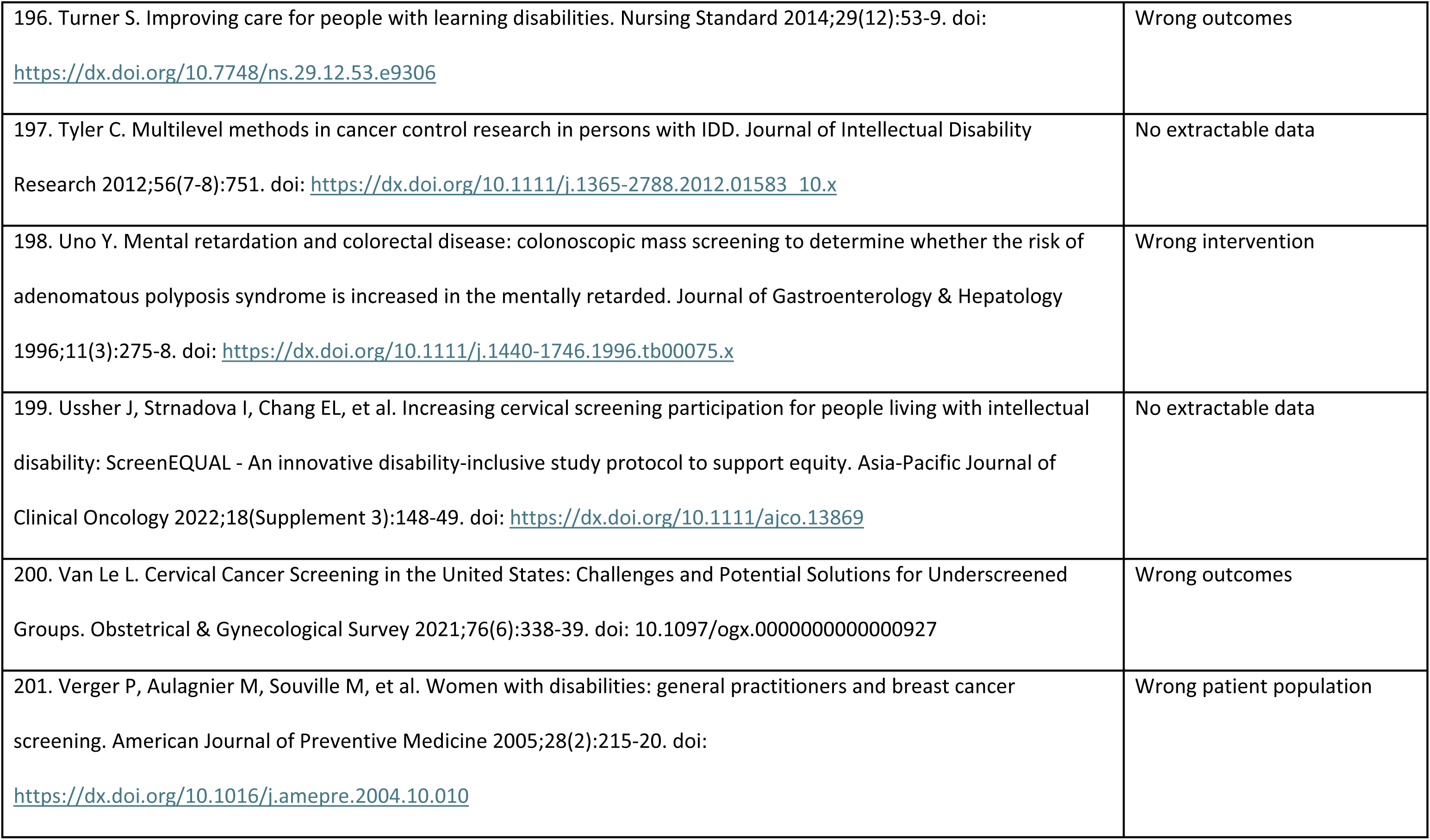
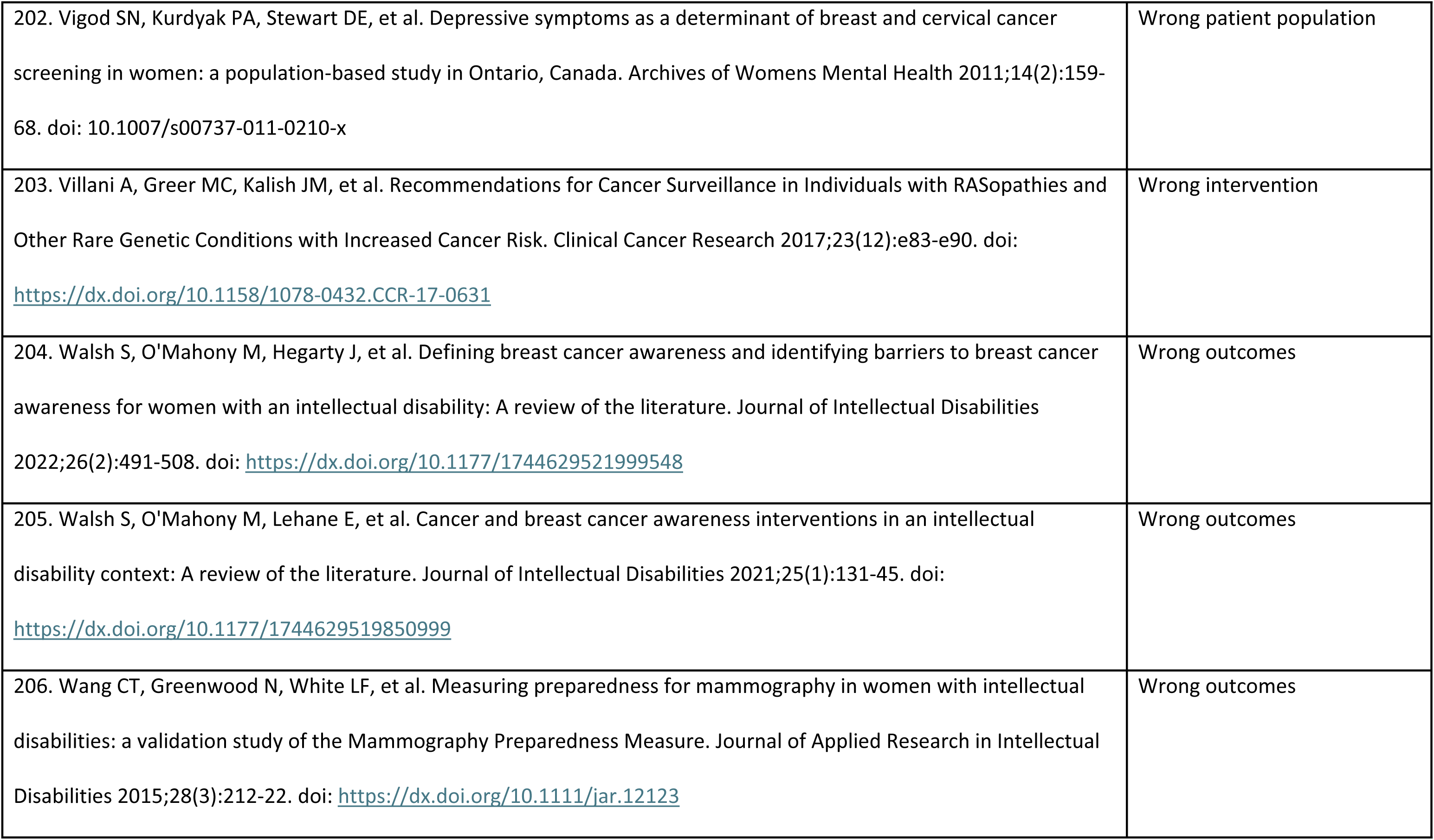
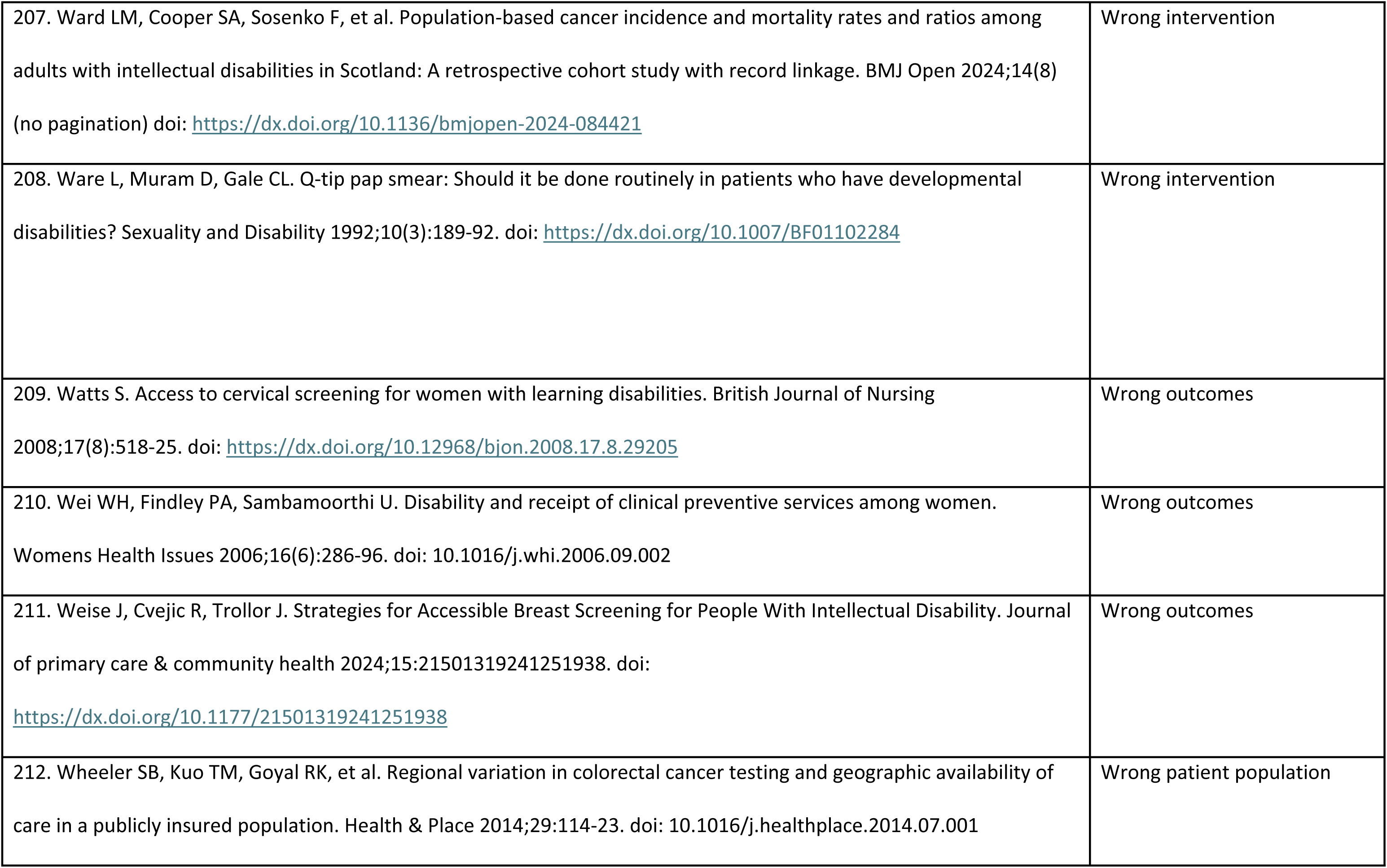
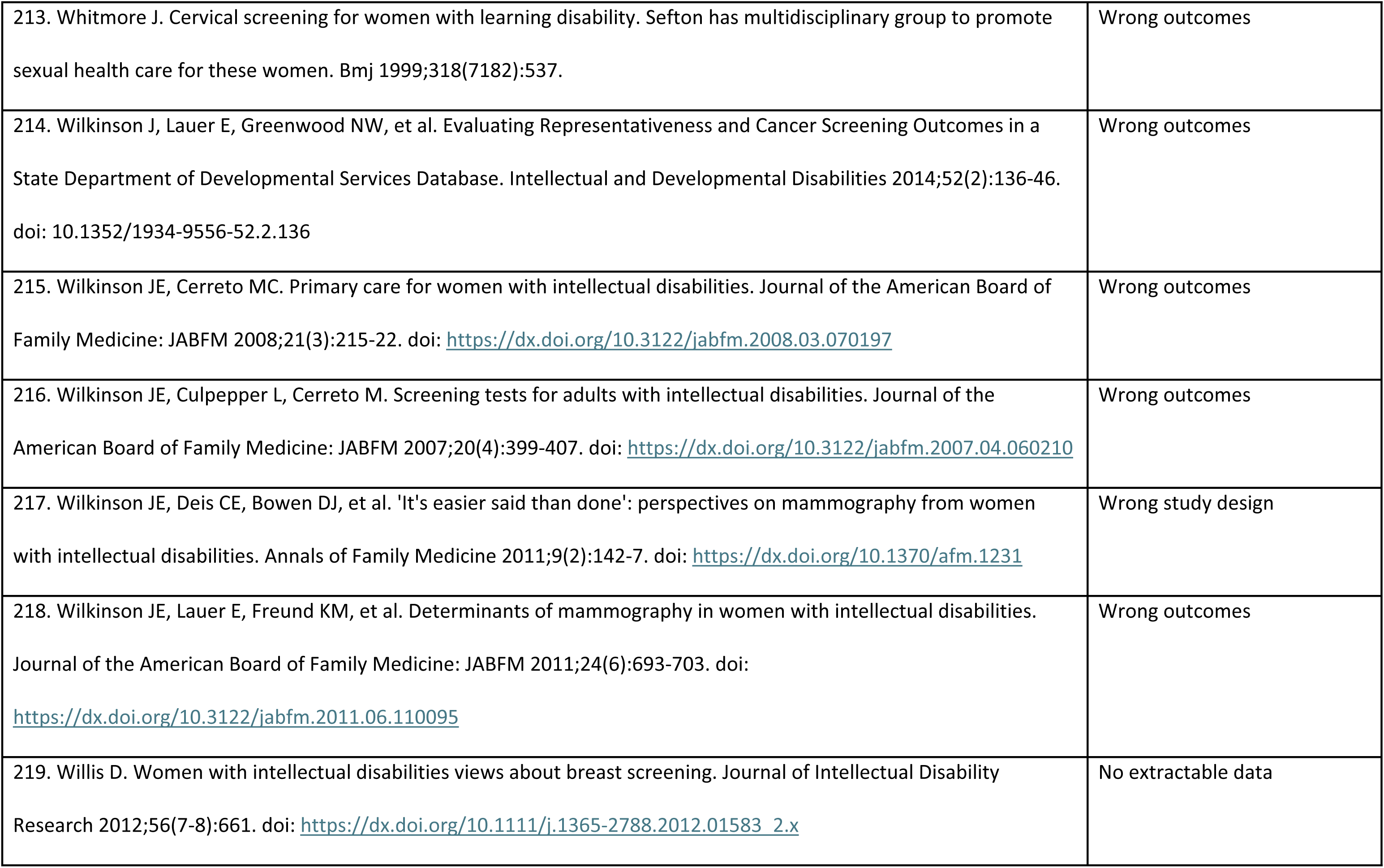
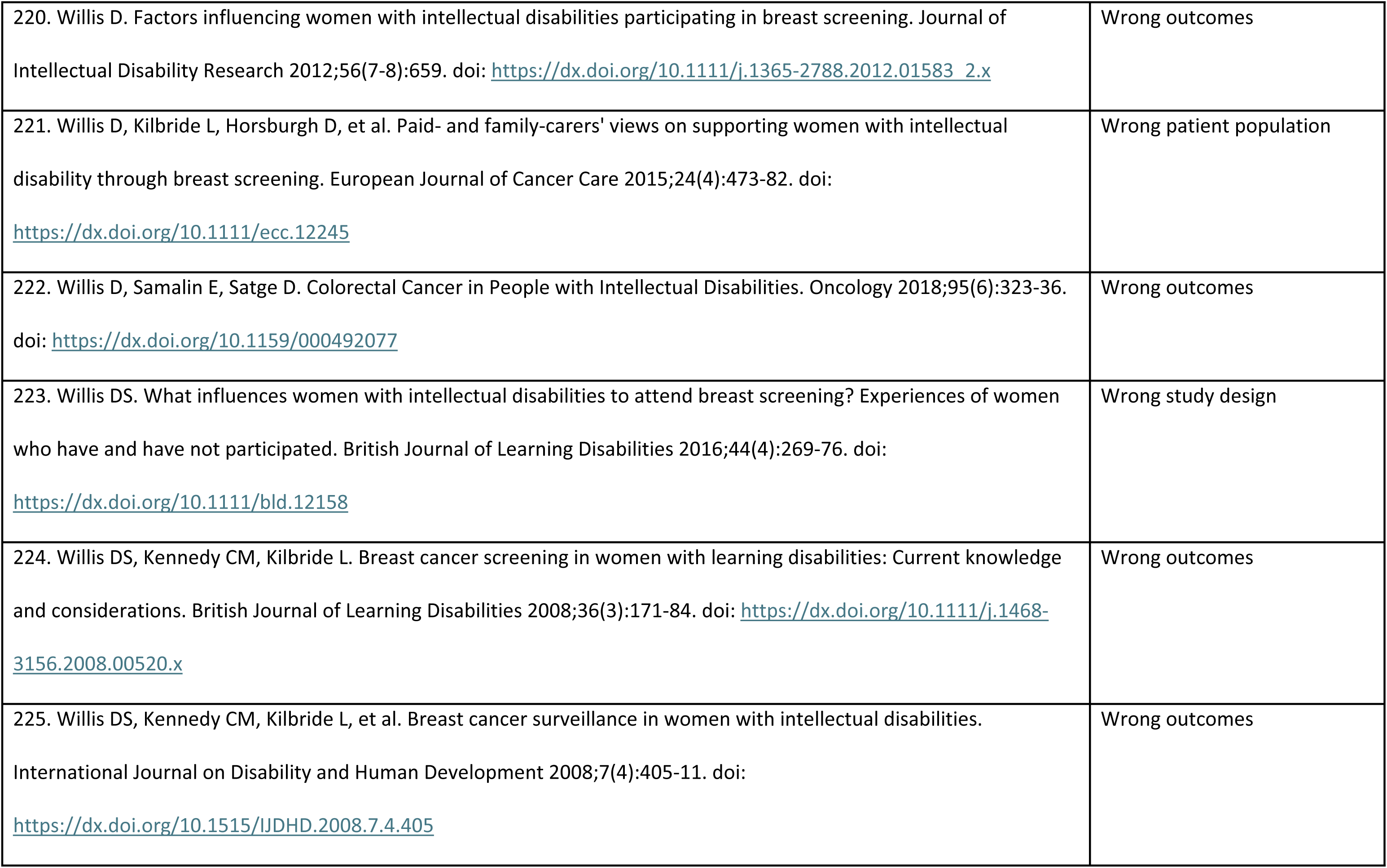
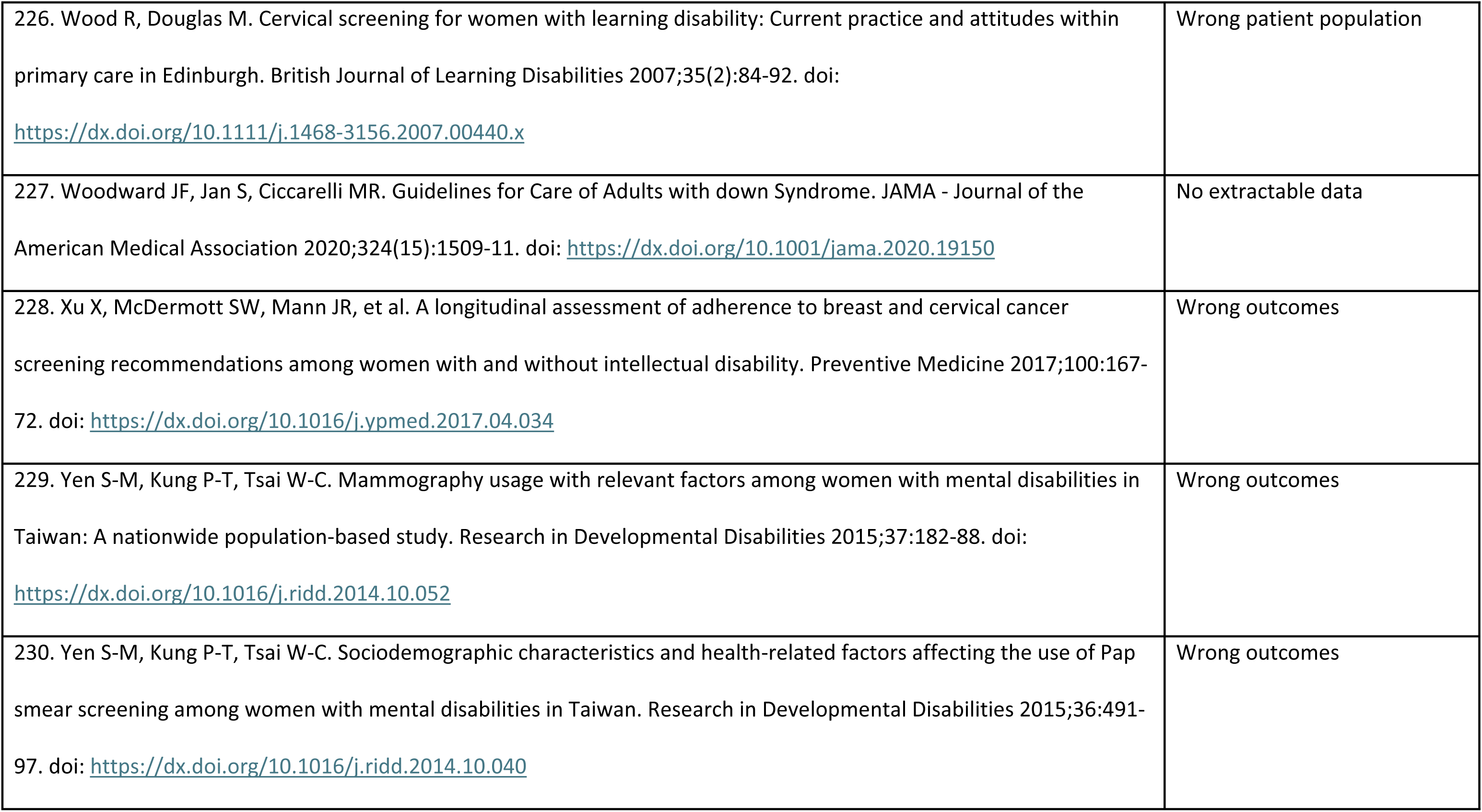
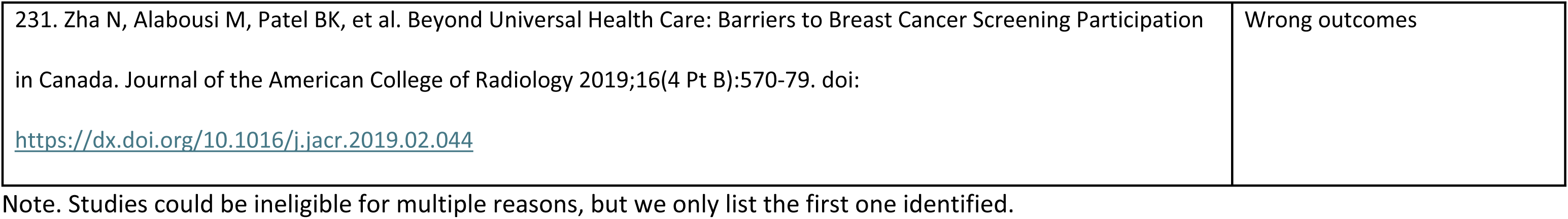

